# Coronary Artery Disease Risk of Familial Hypercholesterolemia Genetic Variants Independent of Historical Cholesterol Exposure

**DOI:** 10.1101/2020.11.12.20230904

**Authors:** Shoa L. Clarke, Catherine Tcheandjieu, Austin Hilliard, Kyung Min Lee, Julie Lynch, Kyong-Mi Chang, Donald Miller, VA Million Veteran Program, Joshua W. Knowles, Christopher O’Donnell, Phil Tsao, Daniel J. Rader, Peter W. Wilson, Yan V. Sun, Michael Gaziano, Themistocles L. Assimes

**Author notes:** **Address for correspondence** Shoa L. Clarke, MD, PhD, 870 Quarry Road, Falk Building, Stanford, CA 94306, Themistocles L. Assimes, MD, PhD, 1070 Arastradero Road, Suite 300, Palo Alto, CA 94304.

## Abstract

**Aims:** Familial hypercholesterolemia (FH) genetic variants confer risk for coronary artery disease (CAD) even after adjusting for low-density lipoprotein cholesterol (LDL-C) levels, using a single measurement. This study evaluated whether multiple historical measures of LDL-C observed in the electronic health record (EHR) can account for the risk associated with FH variants.

**Methods and Results:** We analyzed 418,790 participants in the Million Veteran Program with EHR data spanning up to 15 years prior to and 7 years after enrollment, including ∼6.3 million LDL-C measurements. FH variants in LDLR, *APOB*, and *PCSK9* were identified using a custom genotype array. We implemented a nested case-control design, using incidence density sampling to match etiologic exposure windows and measure CAD risk while adjusting for LDL-C exposure. In a cohort of 23,091 primarily prevalent cases and 230,910 matched controls, FH variants conferred increased risk for CAD (odds ratio: 1.53; 95% confidence interval: 1.24 to 1.89; p: 7.8×10^−5^). Adjusting for mean LDL-C exposure prior to the index date attenuated this risk more than adjusting for a single measurement, but significant risk remained (odds ratio: 1.33; 95% confidence interval: 1.08 to 1.64; p = 8.4×10^−3^). The pattern was also apparent in stratified analyses by sex and ancestry, and we found evidence of an interaction between sex and FH carrier status.

**Conclusion:** The risk associated with FH variants cannot be fully captured by the LDL-C data available in the EHR, even when considering multiple LDL-C measurements spanning more than a decade.

## Introduction

Prevention of coronary artery disease (CAD) through the identification and treatment of risk factors is a cornerstone of primary care and cardiology. In this respect, familial hypercholesterolemia (FH) presents both an opportunity and a challenge. FH is a monogenic disorder that causes elevated low-density lipoprotein cholesterol (LDL-C) from birth, which in turn leads to increased risk for cardiovascular disease. Early identification and treatment of individuals with FH may significantly improve outcomes.^1,2^ However, FH is underdiagnosed and undertreated.^3^ Current practice relies on family history, physical exam, and cholesterol screening to identify patients with FH. Yet, many individuals with FH-causing genetic variants do not meet criteria for the clinical diagnosis of FH,^4^ and adults harboring FH variants may have normal or only mildly elevated LDL-C.^4–6^

Prior studies suggest that FH variants confer risk for CAD that is independent of LDL-C,^5,6^ and FH carriers have increased risk for CAD compared to non-carriers even among individuals with normal LDL-C.^5^ These observations have supported efforts to increase genetic testing for suspected FH.^7^ However, these findings are limited by the incorporation of only a single LDL-C measurement into risk prediction models. In clinical settings, providers often have access to multiple historical LDL-C measurements documented in the medical record. It is unknown whether multiple measurements of LDL-C over time can account for the risk associated with FH variants and potentially negate the added predictive power of genetic testing.

Estimating the risk of FH variants while accounting for repeated measures of LDL-C over many years is challenging given the relatively low prevalence of these variants combined with the small size of most observational cohort studies. However, the maturation of biobanks within large-scale integrated healthcare systems with extensive electronic health records (EHR) provides unprecedent opportunities. Towards this end, we analyzed genetic data and linked EHR-derived outcome data from >400,000 genotyped participants in the Million Veteran Program (MVP)^8^ to test the hypothesis that historical measures of LDL-C over up to twenty years can completely account for the predictive value of FH variant carrier status. Participants provided access to their EHR spanning on average over a decade prior to and up to 7 years after enrollment, during which ∼6.3 million LDL-C measurements were charted in the setting of routine clinical care, and nearly 35,000 cases of CAD accrued.

## Methods

### Study cohort

The MVP cohort has been previously described.^8^ Briefly, United States veterans receiving care at more than 60 Veterans Affairs (VA) Medical Centers across the country have been recruited on an ongoing basis since 2011. DNA from peripheral blood collected at the time of enrolment was used to genotype participants with a custom array, enriched for known pathogenic variants, rare missense variants, indels, and loss-of-function variants.^9^ These data were linked to EHR data, including International Classification of Disease (ICD) diagnosis and procedure codes, Current Procedural Terminology (CPT) codes, prescription data, and clinical laboratory measurements. A majority of EHR-based follow up for MVP participants is presently still captured prior to enrollment, given the VA adopted an EHR over 2 decades ago, and most veterans participating in MVP have received care at the VA over a similar period of time. The VA Central Institutional Review Board approved the MVP study protocol in accordance with the principles outlined in the Declaration of Helsinki. Informed consent was obtained from all participants.

### Study design

The primary outcome of this study was clinically evident CAD, defined by the presence of relevant ICD-9, ICD-10, and CPT codes within the EHR occurring anytime between June 1991 and August 2018. An individual was classified as a case if he or she had ≥1 admission to a VA hospital with discharge diagnosis of acute myocardial infarction (AMI) or ≥1 procedure code for revascularization of the coronary arteries.

We first performed a standard case-control approach in order to provide a direct comparison to prior population studies of FH variant carriers. All unrelated subjects with demographic and EHR data were eligible for this analysis (**Figure 1A**). Controls were defined as individuals without any codes for AMI or revascularization who also did not have any other codes suggestive of coronary disease (**Supplementary material online, Table S1**). Subjects that did not meet criteria to be either a case or a control were considered ambiguous and excluded from the standard case-control analysis.

**Figure 1.**
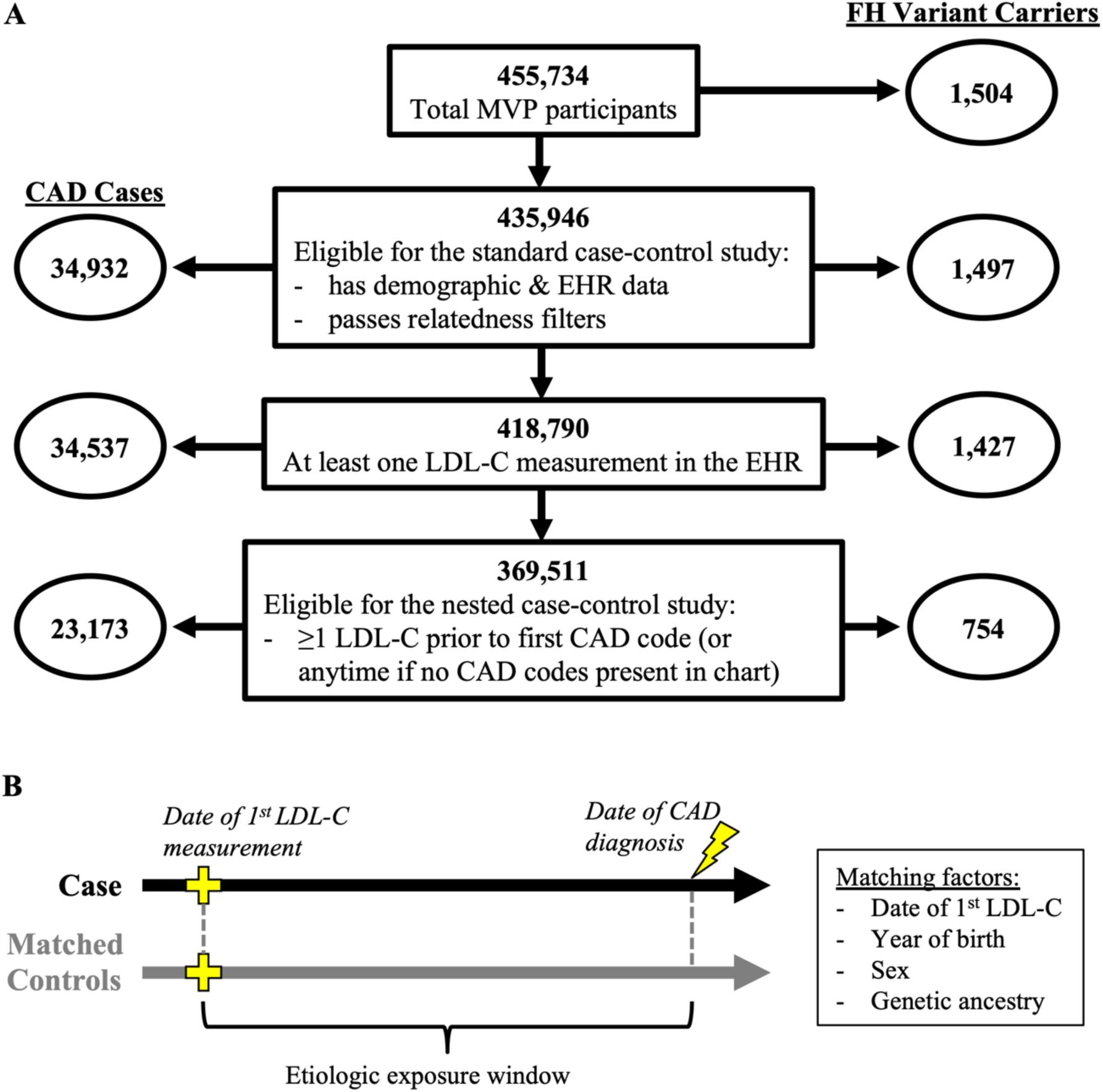
Study Cohort and Study Design. (A) Summary of the study cohort at each stage of analysis. (B) Illustration of the principle of incidence density sampling used to generate matched sets for the nested case-control study. For each case, the index date was set to the date of the first CAD code. Any subject with no CAD codes before or within 1 month after the index date was eligible to serve as a control, and 10 random controls were selected, matching on the date of the first LDL-C measurement, the year of birth, sex, and genetic ancestry. FH = familial hypercholesterolemia. CAD = coronary artery disease.

We then performed a second analysis to measure CAD risk while accounting for longitudinal LDL-C exposure. This analysis used a nested case-control study designed that used the principle of incidence density sampling in order to match the appropriate etiologic exposure window (**Figure 1B**). Cases with at least one LDL-C measurement prior to the first documented CAD code in the EHR were eligible for this study. The date of the first LDL-C was considered time-zero of follow-up and the date of first CAD code that followed was considered the index date. For a given case, any subject with no CAD codes before or within 1 month after the index date was eligible to serve as a control. For each case, 10 random controls were selected, matching on the date of the first LDL-C measurement ± 12 months, the year of birth ± 3 years, sex, and genetic ancestry. We note that time-zero and the index date could fall before the date of enrollment into MVP for many matched case-control sets given the extensive historical EHR data that was available for most participants.

### Familial hypercholesterolemia genetic variant status

Sample collection, genotyping array, and quality control for MVP have been previously described.^9^ Minimac version 4 was used for imputation with the 1000 Genomes phase 3 (version 5) reference haplotypes.^10^

We assessed the *LDLR, APOB*, and *PCSK9* loci for putative FH-causing variants. The start and end coordinates of the 5’ and 3’ UTR for each locus were identified using the human genome hg19 build in the UCSC Genome Browser.^11^ An additional 50 base pairs were added to the ends. Variants were filtered, and heterozygotes were identified using Plink.^12^ Only variants with a minor allele frequency (MAF) < 1% across MVP were considered. In addition to directly genotyped variants, high quality imputed variants were also considered if the ancestry specific R^2^ was greater than 0.8. However, all variants that were ultimately identified as FH variants using the criteria below were directly genotyped.

FH variants were identified using a well validated approach^5,13^ that incorporates 1) ClinVar^14^ annotations of *LDLR, APOB*, and *PCSK9*; 2) predicted loss-of-function variants in *LDLR*; and 3) predicted pathogenic missense variants in *LDLR*. Annovar^15^ was used to annotate variants based on ClinVar (version 20190305) classification and dbNSFP (version 3.3) annotations.^16^ *LDLR, APOB*, and *PCSK9* variants that were classified by ClinVar as pathogenic or likely pathogenic for FH were kept as FH variants. Loss-of-function variants in *APOB* and *PCSK9* that were labeled as pathogenic for FH in ClinVar were excluded, as such variant are typically associated with hypocholesterolemia rather than hypercholesterolemia. Through manual review, we found that these annotations likely represent errors in ClinVar version 20190305. *LDLR* loss-of-function variants were predicted using the LOFTEE plugin to the Variants Effect Predictor tool. Predicted pathogenic missense variants in *LDLR* were defined as variants predicted to be pathogenic by all five of five tools (SIFT, PolyPhen-HumDiv, PolyPhen-HumVar, MutationTaster, and LRT). No *LDLR* predicted loss-of-function variant or predicted pathogenic missense variant was classified as benign or possibly benign by ClinVar. Lastly, a rare *APOB* missense variant (hg19 chr2:21229160:C:T) that was marked as “conflicting interpretations of pathogenicity” in ClinVar was included in our analyses as an FH variant based on prior observations^17^ and results presented here. A subject was identified as a heterozygous carrier of an FH variant if the estimated probability of being a heterozygote for the given variant was ≥0.95, as calculated by Plink.^12^

### Ancestry

An individual’s ancestry was determined using the HARE algorithm,^18^ with each participant being assigned to one of five mutually exclusive ancestry groups: non-Hispanic African, non-Hispanic-Asian, non-Hispanic European, Hispanic, or unclassified. The unclassified category represents highly admixed individuals who could not be clearly classified into any one ancestry group that was congruent with their self-report.

### Relatedness

We used KING version 2.0 to identify individuals who were first, second, or third-degree relatives to one another.^19^ For each pair of relatives, we removed one individual from the cohort. When possible, we preferentially retained individuals carrying FH variants.

### Low-density lipoprotein cholesterol measures

Laboratory test results generated between December 1998 and October 2018 were extracted from the EHR. Test names reflecting blood LDL-C levels were adjudicated and harmonized by two clinicians. Only values with the specified unit of mg/dL were retained, and values less than 20 mg/dL or greater than 600 mg/dL were excluded as outliers. The VA prescription database was also adjudicated and harmonized by two clinicians to identify drug names that correspond to statins. Included statins were atorvastatin, cerivastatin, fluvastatin, lovastatin, pitavastatin, pravastatin, rosuvastatin, and simvastatin. In addition, combination drugs that contained any of these statins were included. We used the medication dispense date from the VA pharmacy to identify receipt of statin prescriptions. An individual was considered on statin from the date of dispensation through the length of the days-supply plus a buffer of up to an additional 30-days. LDL-C measurements occurring during statin treatment were divided by 0.7 to adjust for statin use.^5,6,20^

We derived three different LDL-C exposures using multiple measures of LDL-C available to us: first LDL-C was defined as the first LDL-C within a time window; max LDL-C was defined by the highest measure within a time window; mean LDL-C was defined as the mean of all LDL-C measures available within a time window. These LDL-C exposure metrics were calculated using statin-adjusted values where appropriate.

### Traditional risk factors

Traditional risk factors for CAD considered in our analyses included hypertension, diabetes, and tobacco use. These risk factors were identified using diagnosis codes (**Supplementary material online, Table S2**) available between June 1991 and August 2018. A risk factor was considered present if two or more codes for a given risk factor were identified.

### Statistical analysis

We generated density distributions of the three statin-adjusted LDL-C measures (first, maximum, and mean) by FH carrier status using the full set of LDL-C data for each individual, and receiver operating characteristic curves for predicting the presence of an FH variant using each of the same three LDL-C measures, accounting for age at the time of the measurement. Descriptive statistics for the nested case-control cohort were calculated using the EHR data up until the index date for each matched set. For all other contexts, descriptive statistics were calculated using the full span of data in the EHR.

We calculated the odds ratio (OR) for CAD using unconditional multivariate logistic regression for both analyses. The regression for the standard case-control analysis included adjustment for year of birth, sex, genetic ancestry, hypertension, diabetes, tobacco use, and statin use. Cases were considered to be hypertensive, diabetic, and users of tobacco only if at least one of the two qualifying ICD codes for these derived variables occurred prior to the first CAD code. Statin use was considered present for a case if there was at least one statin prescription prior to the first CAD code. When further adjusting for LDL-C, we used the first documented LDL-C measure. Individuals whose first available LDL-C measurement occurred after the first CAD code were included in this analysis.

The regression for our nested case-control analysis was adjusted for all matching factors (age, sex, genetic ancestry, and date of first LDL-C)^21^ as well as hypertension, diabetes, tobacco use, statin use, and the number of LDL-C measurements within the etiologic exposure window. For both cases and controls, hypertension, diabetes, tobacco use, and/or statin use was considered to be present if first documented prior to the index date. Adjustments for LDL-C exposure were based on LDL-C measurements occurring within the etiologic exposure window (date of first LDL-C to index date). Sex and ancestry were each assessed for interaction effects with FH variant carrier status by adding an interaction term to the regression. Lastly, we conducted sensitivity analyses to determine if the effect of LDL-C adjustment on the risk of CAD for the presence of an FH variant was modified by the length of time and number of LDL-C measurements between the first and the last LDL-C measures within an etiologic exposure window. For these analyses, we created new matched sets not only using the matching factors already described but also matching on the defined minimal number of LDL-C measurements and minimal span of time covered by these measurements.

Statistical analyses were performed using R version 3.5.1 (R Foundation, Vienna, Austria).

## Results

### Prevalence of FH variants in the MVP population

We identified 55 FH variants (51 *LDLR*, 2 *APOB*, 2 *PCSK9*) among 455,734 genotyped MVP participants (**Supplementary material online, Table S3**). FH variants were defined by 1) ClinVar annotations of *LDLR, APOB* and *PCSK9*; 2) predicted loss-of-function variants in *LDLR*; and 3) predicted pathogenic missense variants in *LDLR*. Additionally, we assessed two missense variants in *APOB* that were previously found to be associated with severe hypercholesterolemia in MVP ^17^ but were labeled as “uncertain” or “conflicting evidence” in ClinVar. We found that one of these variants was strongly associated with CAD (**Supplementary material online, Table S4**), and thus we chose to keep it in our analysis as an FH variant. All identified FH variants were directly genotyped. In total, we found 1,504 heterozygous carriers of FH variants, for an approximate prevalence of 1 in 303. The European genetic ancestry group had the highest prevalence of FH variant carriers, and the Asian genetic ancestry group had the lowest prevalence, but the prevalence was similar across all ancestry groups (**Table 1**). After excluding individuals with missing demographic data and filtering for relatedness, we were left with 435,946 unrelated individuals, including 1,497 FH carriers (**Figure 1A**).

**Table 1.** Prevalence of FH variant carriers in the Million Veteran Program.

|  | Genetic ancestry group |  |  |  |  |  |
| --- | --- | --- | --- | --- | --- | --- |
|  | All | African | Asian | European | Hispanic | Unclassified |
| n | 455,734 | 87,163 | 4,553 | 318,694 | 34,151 | 11,173 |
| FH variant carriers | 1,504 | 258 | 11 | 1,095* | 111 | 29 |
| *LDLR* LoF | 165 | 20 | 3 | 130 | 10 | 2 |
| *LDLR* missense | 944 | 222 | 6 | 606 | 91 | 19 |
| *APOB* | 383 | 16 | 2 | 349 | 8 | 8 |
| *PCSK9* | 13 | 0 | 0 | 11 | 2 | 0 |
| Prevalence | 1:303 | 1:338 | 1:414 | 1:291 | 1:308 | 1:385 |
| (95% CI) | (1:288-319) | (1:301-385) | (1:260-1,010) | (1:275-309) | (1:259-378) | (1:283-605) |

\* One individual was found to be a carrier of both an *LDLR* missense variant and an *APOB* variant.
LoF = loss of function. CI = confidence interval.

### LDL-C metrics and association with FH carrier status

The majority of genotyped participants (418,790 or 96.1%) had at least one LDL-C measurement in the EHR, and the median number of LDL-C measurements per individual was 12 (interquartile range 6-21). In total, ∼6.3 million LDL-C measurements were used in this study. LDL-C measurements were correlated with ∼5.3 million recorded statin prescriptions, and LDL-C values were adjusted for statin use where appropriate.

MVP participants with FH variants showed a wide range of LDL-C values (**Figure 2**). The prevalence of FH variant carriers among subjects with severe hypercholesterolemia (LDL-C ≥ 5 mmol/L) depended on which LDL-C metric was used to define severe hypercholesterolemia. If only considering the first available LDL-C, the prevalence was 1.4%. When considering all subjects who ever demonstrated severe hypercholesterolemia (i.e. max LDL-C ≥ 5 mmol/L), the prevalence of FH variant carriers decreased to 0.7%. Alternatively, when using the mean of all observed LDL-C values to define severe hypercholesterolemia, the prevalence of FH variant carriers increased to 3.5%. Using a more stringent LDL-C threshold of ≥ 6 mmol/L, the prevalence of FH carriers for each LDL-C metric was 3.1%, 1.1%, and 8.8% respectively (**Table 2**). In general, however, LDL-C alone offers only modest discriminatory power for predicting the presence or absence of FH variants, with mean LDL-C performing better than first or maximum observed LDL-C (**Supplementary material online, Figure S1**).

**Table 2.** Prevalence of FH variant carriers by LDL-C level, defined by the first available, the maximum observed, or the mean of all measures.

|  | n | FH variant carriers (%) |
| --- | --- | --- |
| **First LDL-C (mmol/L)** |  |  |
| <3 | 203,772 | 465 (0.2) |
| 3-4 | 141,951 | 411 (0.3) |
| 4-5 | 56,836 | 317 (0.6) |
| 5-6 | 12,767 | 125 (1.0) |
| ≥6 | 3,464 | 109 (3.1) |
| **Max LDL-C (mmol/L)** |  |  |
| <3 | 78,911 | 144 (0.2) |
| 3-4 | 134,427 | 295 (0.2) |
| 4-5 | 109,353 | 319 (0.3) |
| 5-6 | 57,394 | 255 (0.4) |
| ≥6 | 38,705 | 414 (1.1) |
| **Mean LDL-C (mmol/L)** |  |  |
| <3 | 218,118 | 430 (0.2) |
| 3-4 | 158,680 | 526 (0.3) |
| 4-5 | 37,367 | 308 (0.8) |
| 5-6 | 4,060 | 113 (2.8) |
| ≥6 | 565 | 50 (8.8) |

LDL-C = low-density lipoprotein cholesterol
FH = familial hypercholesterolemia

**Table 3.** Characteristic of the nested case-control cohort.

| Characteristic | CAD cases | Matched Controls |
| --- | --- | --- |
| Demographics |  |  |
| n | 23,091 | 230,910 |
| Male | 22,497 (97.4) | 224,870 (97.4) |
| Age at enrollment (years) | 66.3 ± 9.1 | 66.3 ± 9.0 |
| Ancestry |  |  |
| African | 3,620 (15.7) | 36,200 (15.7) |
| Asian | 144 (0.6) | 1,440 (0.6) |
| European | 17,553 (76.0) | 175,530 (76.0) |
| Hispanic | 1,434 (6.2) | 14,340 (6.2) |
| Unclassified | 340 (1.5) | 3,400 (1.5) |
| Lipid Data |  |  |
| Age at first LDL-C (years) | 57.3 ± 9.0 | 57.2 ± 9.0 |
| First LDL-C to index date (years) | 5.7 ± 4.6 | 5.7 ± 4.5 |
| LDL-C (mmol/L) |  |  |
| First | 3.4 ± 1.1 | 3.2 ± 1.0 |
| Maximum prior to index date | 4.2 ± 1.4 | 3.9 ± 1.3 |
| Mean prior to index date | 3.4 ± 0.9 | 3.2 ± 0.9 |
| Medical history |  |  |
| Hypertension |  |  |
| prior to first LDL-C | 11,447 (49.6) | 93,759 (40.6) |
| prior to index date | 17,938 (77.7) | 148,201 (64.2) |
| Diabetes |  |  |
| prior to first LDL-C | 5,634 (24.4) | 33,379 (14.5) |
| prior to index date | 9,438 (40.9) | 61,175 (26.5) |
| Tobacco |  |  |
| prior to first LDL-C | 4,115 (17.8) | 32,561 (14.1) |
| prior to index date | 8,320 (36.0) | 62,993 (27.3) |
| Statin use |  |  |
| prior to first LDL-C | 3,882 (16.8) | 29,076 (12.6) |
| prior to index date | 14,644 (63.4) | 113,023 (48.9) |
| FH variant carrier | 103 (0.4) | 651 (0.3) |
| Case type |  |  |
| Prevalent cases | 17,642 (76.4) | NA |
| Index date to enrollment (years) | 5.7 (4.1) | NA |
| Incident cases | 5,449 (23.6) | NA |
| Enrollment to index date (years) | 2.0 (1.5) | NA |

Values are n (%) or mean ± SD.
CAD = coronary artery disease
LDL-C = low-density lipoprotein cholesterol
FH = familial hypercholesterolemia

**Figure 2.**
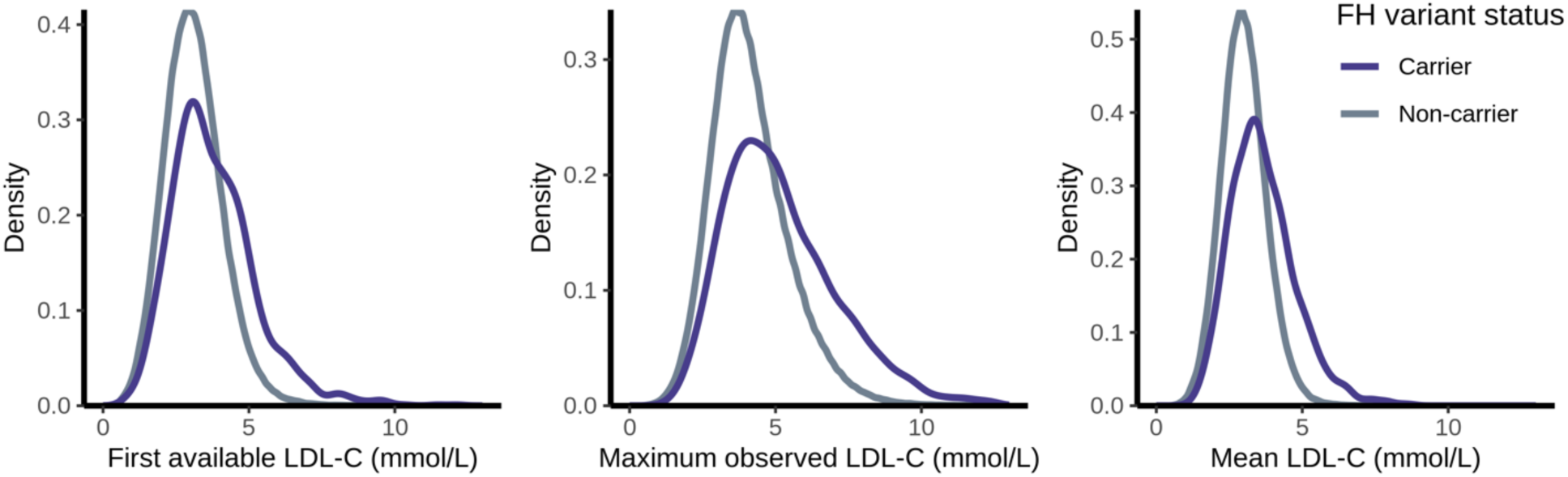
Observed LDL-C in FH Variant Carriers and Non-carriers. Density distributions of the first, maximum, and mean low-density lipoprotein cholesterol (LDL-C) measurements observed in the electronic health record (EHR) for individuals with and without familial hypercholesterolemia (FH) genetic variants.

### FH genetic variants, LDL-C exposure, and risk for CAD

We first conducted a standard case-control study of CAD in order to provide comparison to prior sequencing-based population studies of FH variant carriers.^4,6^ We identified 34,932 CAD cases (**Figure 1A**). A majority of cases (29,300; 84%) were prevalent at the time of enrollment with a mean time from first CAD code to enrollment of 7.6 ± 4.9 years. For incident cases occurring after enrollment, the mean time from enrollment to the date of the first CAD code was 2.0 ± 1.5 years. We compared these cases to 291,408 controls defined as having no codes suggestive of CAD documented across the full span of EHR data. All traditional risk factors were more prevalent among cases compared to controls, and individuals who could not be classified as either case or control tended to have risk factor prevalence intermediate to the cases and controls (**Supplementary material online, Table S5**). FH carriers had a 1.7-fold (95% CI: 1.4 to 2.0; p = 1.9×10^−9^) increased odds for CAD compared to noncarriers and a 3.0-fold (95% CI: 1.7 to 5.0; p = 5.4×10^−5^) increased odds for premature CAD (male <55 and female <65), consistent with other population studies^4,6^ (**Supplementary material online, Figure S2**). When adjusting for LDL-C using the first available measurement, the risk associated with FH variants attenuated but remained significant. For all CAD, the odds ratio attenuated to 1.4 (95% CI: 1.2 to 1.6; p = 3.2×10^−4^). For premature CAD, the odds ratio attenuated to 2.1 (95% CI: 1.2 to 3.7; p = 7.9×10^−3^).

We next conducted a nested case-control study designed to measure the risk of CAD while adjusting for LDL-C exposure over the appropriate etiologic window. To do so, cases were restricted to those with LDL-C measurements available prior to the first diagnosis of CAD (n = 23,173). We found most cases to have substantial prior LDL-C data, with the median number of prior measurements being 6 (interquartile range 2-12) and the median span between the first measurement and the most recent measurement prior to the CAD diagnosis being 49 months (interquartile range 12-100). When stratifying cases by FH carrier status, we found both carriers and non-carriers had similarly extensive prior LDL-C data (**Supplementary material online, Figure S3**). Controls were selected using incidence density sampling (10 controls per case), matching sex, year of birth, ancestry, and date of first LDL-C measurement (**Figure 1B**). In total, 23,091 cases (99.6%) were successfully matched to 10 controls (**Table 2**). The odds of CAD for FH variant carriers compared to non-carriers among matched case-control sets was 1.53-fold higher (95% CI: 1.24 to 1.89; p = 7.8×10^−5^). When adding an adjustment for the first LDL-C measurement, the maximum observed LDL-C prior to the index date, or the mean LDL-C prior to the index date, the odds ratio progressively attenuated, but the risk of FH variants remained significant (**Figure 3**). Consistent with these findings, we found an analogous increase in the magnitude of the odds ratio associated with CAD for each of the three LDL-C measures, with mean LDL-C contributing the highest risk with the most precision (**Supplementary material online, Table S6**). We also observed the same pattern of incomplete attenuation when analyzing matched sets of incident cases only (**Supplementary material online, Figure S4**).

**Figure 3.**
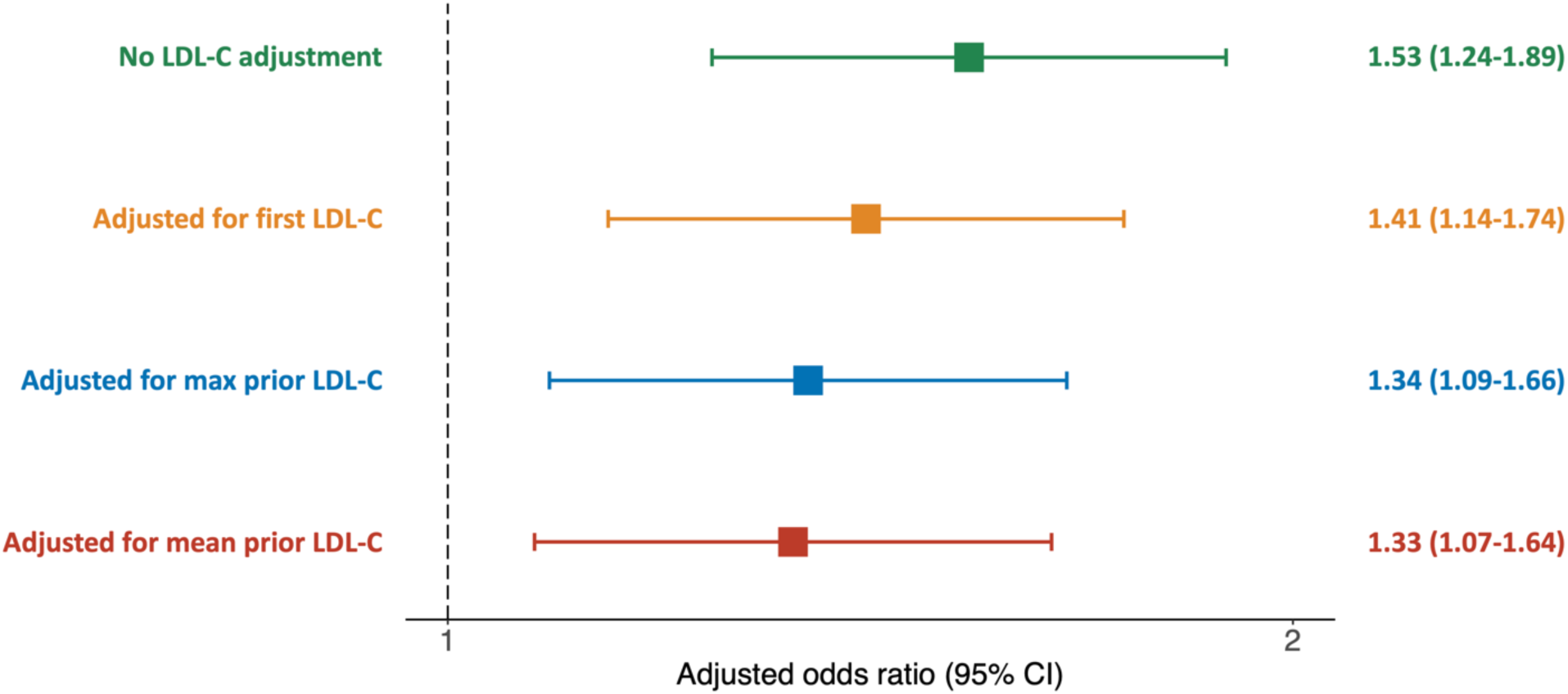
Association between FH Variants and CAD with Adjustments for Historical LDL-C Exposure. Risk of coronary artery disease (CAD) associated with familial hypercholesterolemia (FH) genetic variants. Adjustment for the first, maximum observed, or mean low-density lipoprotein cholesterol (LDL-C) prior to the index date does not fully attenuate the risk associated with FH. Odds ratios were estimated using logistic regression, adjusting for the nested case-control matching factors, tobacco use, hypertension, diabetes, statin prescription, and number of LDL-C measurements.

We next tested whether sex modified the effects of FH variants on CAD risk. Using logistic regression with male non-carriers as the reference group, we found a significant interaction between female sex and FH carrier status (p = 0.03). The interaction remained significant with adjustments for LDL-C (**Figure 4A**). Stratified analyses showed that female FH carriers were at a 3.65-fold (CI: 1.51 to 8.84; p = 4.1×10^−3^) higher risk compared to female non-carriers, while male carriers had a 1.46-fold higher (CI: 1.17 to 1.82; p = 7.0×10^−4^) risk compared to male non-carriers (**Figure 4A**). Several differences between male and female subjects in MVP may contribute to this pattern (**Supplementary material online, Table S7**). Importantly, female subjects were younger than male subjects on average. We also found that female FH carriers tend to have higher LDL-C than male FH carriers, while female and male non-carriers have relatively similar LDL-C. Statin use and CAD risk factors are less prevalent among female subjects compared to males.

**Figure 4.**
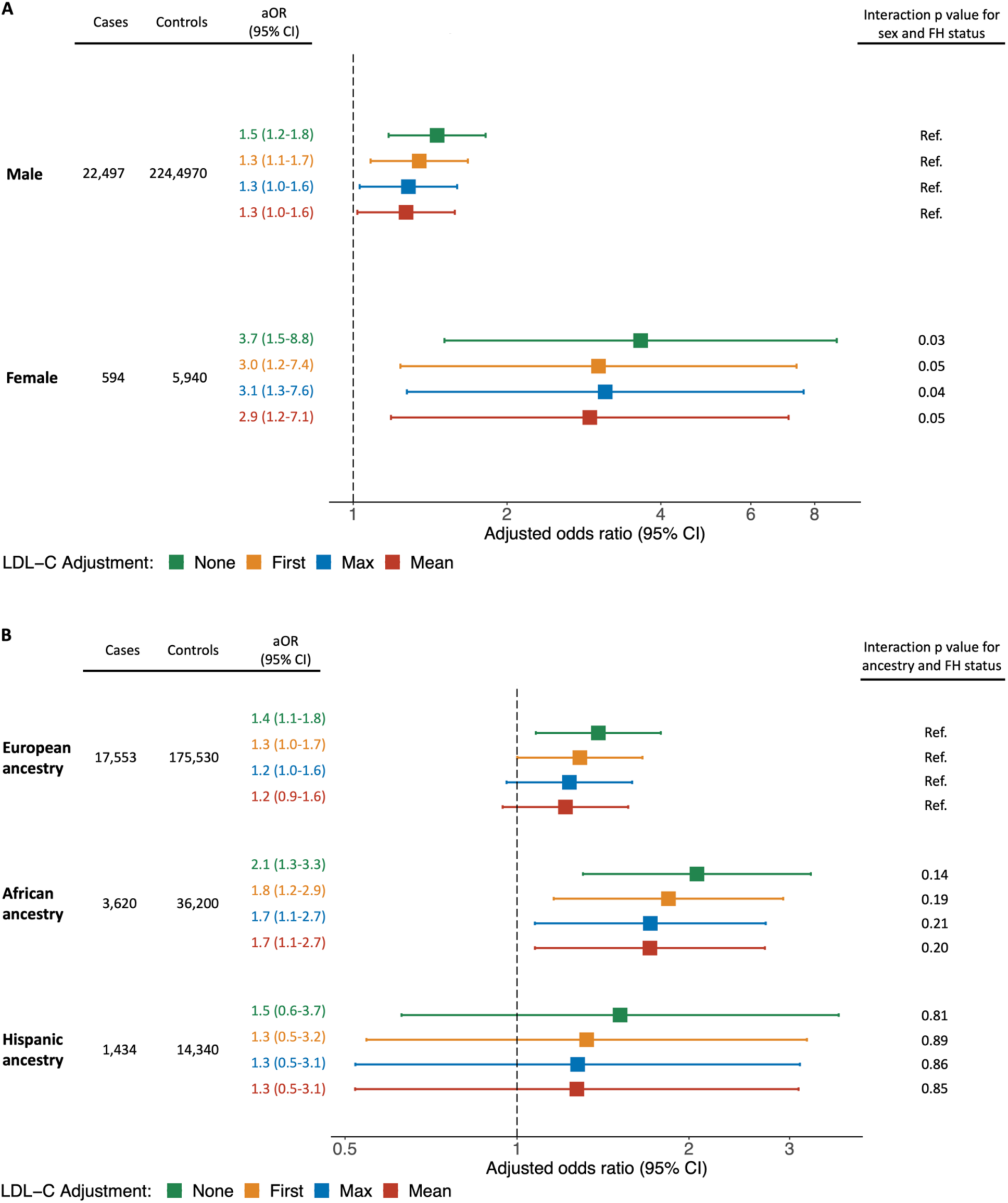
Evaluation of Sex and Ancestry Risk Modifying Effects on FH Carrier Status and CAD. Risk of coronary artery disease (CAD) associated with familial hypercholesterolemia (FH) genetic variants, stratified by sex (A) and by ancestry (B). To the right are p values for test of interaction by logistic regression in the non-stratified data, where “Ref.” denotes the reference group. Odds ratios were estimated using logistic regression, adjusting for the nested case-control matching factors, tobacco use, hypertension, diabetes, statin prescription, and number of LDL-C measurements.

We also tested for interactions between FH variant carrier status and genetic ancestry. Using non-carriers with European ancestry as a reference, we did not find a statistically significant interaction between ancestry and FH carrier status. Though, we saw a trend towards significance in relation to African ancestry (**Figure 4B**). Stratified analyses showed that within the African ancestry group, carriers had a 2.06-fold (CI: 1.30 to 3.27; p = 2.0×10^−3^) higher CAD risk, and within the European ancestry group, carriers had a 1.39-fold higher CAD risk (CI: 1.08 to 1.79; p = 1.1×10^−2^) (**Figure 4B**). Notably, MVP subjects with African ancestry tend to be younger than those with European ancestry (**Supplementary material online, Table S8**).

Lastly, we sought to determine if the pattern we observed in this study – incomplete attenuation of CAD risk with adjustments for LDL-C exposure – was driven primarily by subjects with the limited historical LDL-C data. We therefore generated and assessed matched sets of subjects with extensive LDL-C data. In a matched cohort requiring ≥5 LDL-C measures spanning ≥5 years prior to the index date (9,786 cases, 97,860 controls) and in a matched cohort requiring ≥10 LDL-C measures spanning ≥10 years (3,615 cases, 36,150 controls), we did not observe any notable differences in the degree of attenuation of the adjusted odds ratio for FH carrier status (**Supplementary material online, Table S9-10**).

## Discussion

In this study, we assessed the relationship between FH variants, longitudinal LDL-C exposure, and CAD risk. We found carriers of FH variants in the MVP cohort to have a wide distribution of LDL-C values, in line with other population studies.^4,6^ The risk associated with FH variants in the MVP cohort was comparable to other studies, when using a standard case-control design (**Supplementary material online, Figure S2**). This risk was independent of LDL-C when considering a single measurement. However, this approach does not allow for adjustment of multiple LDL-C measurements over time which more effectively captures the lifetime differences in LDL-C exposure. To address this issue, we adopted a nested case-control design and carefully matched the etiologic exposure window of controls to cases using the principal of incidence density sampling. Using extensive EHR data, we then showed that adjusting for longitudinal LDL-C exposure using multiple measurements does not fully attenuate the risk associated with FH variants, even among the subset of individuals with ≥10 LDL-C measurements spanning more than a decade.

We also found evidence of a modification of effect of FH variants by sex. Among female subjects, the CAD risk associated with FH variants was higher with and without LDL-C adjustment. This difference may be accounted for by less survival bias. Female subjects in MVP tend to be younger than male subjects and tend to have fewer risk factors. Moreover, premature mortality due to cardiovascular disease is less common in women compared to men.^22^ Other sex differences may also contribute. For example, across childhood and adolescence, untreated girls with FH demonstrate consistently higher LDL-C levels than untreated boys,^23^ and adult women with FH may be undertreated compared to men.^24^ We observed patterns in MVP consistent with these prior findings, but additional studies are needed to better understand sex-differences while accounting for several potential confounders.

An important strength of MVP is the genetic diversity, which is more reflective of the U.S. population than European biobanks. In this study, we were sufficiently powered to analyze the impact of FH variants within a cohort of subjects of non-Hispanic African ancestry. We found that FH variants conferred greater CAD risk among subjects of African ancestry compared to subjects of non-Hispanic European ancestry. We believe that this difference likely reflects selection biases that occur with stratification. However, this difference may also reflect racial disparities in the treatment of FH. For example, in an analysis of clinical FH and self-reported race in the CASCADE-FH registry, U.S. Blacks were more likely to be undertreated compared to whites.^24^ In our cohort, statin use among FH carriers of African and European ancestry was similar (**Supplementary material online, Table S8)**, but additional work is needed to assess timing and adequacy of treatment.

In sum, our observations support the notion that genetic testing adds important predictive value to standard clinical assessment, even when longitudinal LDL-C measures are considered. This finding is consistent with a recently proposed framework that recommends both LDL-C measurement and genetic assessment to identify the highest risk patients.^25^ Our study suggests that among adult patients, typical LDL-C monitoring does not optimally stratify subjects by their lifelong exposure to LDL-C (**Take home figure**). The cholesterol exposure pattern of FH carriers versus non-carriers is most distinct during childhood.^26^ We hypothesize that much of the excess risk associated with FH variants accumulates during childhood and early adulthood, a time when a majority are not treated. Thus, adult FH carriers and non-carriers who demonstrate similar patterns of LDL-C may have already separated their risk trajectories in the decades prior to LDL-C monitoring.

Pediatric guidelines recommend screening LDL-C in children to identify FH early in life.^27^ It is possible that if childhood LDL-C data were available, adjustment for LDL-C exposure over a greater fraction of one’s lifetime may supplant the predictive power of FH variants. However, evaluation of lifelong LDL-C measurements is not currently feasible in most clinical settings, whereas genetic testing is rapidly becoming widely available.

The cost effectiveness of genetic testing for FH remains a debate. Cascade screening is one cost-effective strategy,^28^ but it is underutilized in the United States.^29^ Universal screening may ultimately prove cost-effective when considering the possibility of simultaneously testing for actionable genetic variants across multiple syndromes. For example, ∼1% of UK Biobank subjects harbor pathogenic variants for FH, hereditary breast or ovarian cancer syndrome, or Lynch syndrome.^30^ As genetic testing becomes more informative for a wider spectrum of diseases, and as the cost continues to decline, we expect genetic risk assessment to become an integral part of primary prevention. The existence of effective, safe, and inexpensive primary prevention strategies such as lifestyle counseling and statins affords CAD a major advantage in this respect. Efforts are underway within MVP to implement return of actionable results to research participants, and the presence of an FH variant is one such actionable result being explored.

## Study Limitations

An important limitation of our study is that the majority of the CAD cases are prevalent occurring up to 20 years prior to enrollment. Our risk estimates therefore suffer from survivor bias. Moreover, MVP participants tend to be older at enrollment and have more CAD risk factors when compared to other biobanks. This difference may further enhance survivor bias. Thus, our study likely underestimates the risk of FH variants. However, underestimating the risk of FH is not expected to alter our main conclusion regarding patterns of risk attenuation.

A second limitation of our study is the use of a genotyping array rather than gene sequencing to identify FH variants. Although the MVP array is designed to detect rare protein-altering variants and known disease-causing variants, we expect to miss some rare variants that would be identified with sequencing. The estimated prevalence of FH carriers in a population varies significantly depending on sampling methods, sequencing methods, and bioinformatic analyses.^4,6,31^ Based on prior U.S. data^4^ as well as a recent global meta-analysis,^32^ we may reasonably estimate the expected prevalence of FH variant carriers in our cohort to be 1 in ∼250-300. Thus, we have likely missed only a small number of FH carriers. Furthermore, when using the same study design, our risk estimates for CAD are consistent with other population studies that identified FH carrier by sequencing (**Supplementary material online, Figure S2**).

Finally, the MVP cohort is predominantly male. Although we were reasonably powered to perform sex-stratified analysis, our risk estimates are less precise in women due to a small sample size. Larger studies of FH among women are needed to confirm our findings and to better understand potential sex differences.

## Conclusions

FH genetic variants confer significant risk for CAD. LDL-C exposure defined by serial longitudinal measurements in the EHR can account for some but not all of this risk. The residual risk associated of FH variants reflects the limitations of clinical phenotyping for capturing genetic risk. Whereas FH variants impact LDL-C exposure continuously throughout life, clinical measurements of LDL-C can only sample a fraction of this exposure. Genetic testing improves identification of high-risk patients.

## Data Availability

Individual data cannot be shared publicly according to the Data Access Policy of the Million Veteran Program in the VA Office of R&D in Veterans Health Administration. Requests for data access be submitted to Dr. Jennifer Moser PhD at the Genomic Medicine & Million Veteran Program in the VA Office of R&D.

## Funding

This work was supported by funding from the Department of Veterans Affairs Office of Research and Development, Million Veteran Program Grants 2I01BX003362 and 1I01BX004821. This publication does not represent the views of the Department of Veteran Affairs or the United States Government.

### Acknowledgements

We greatly appreciate the participation of the Million Veterans Program participants and the support of the Million Veterans Program staff.

---

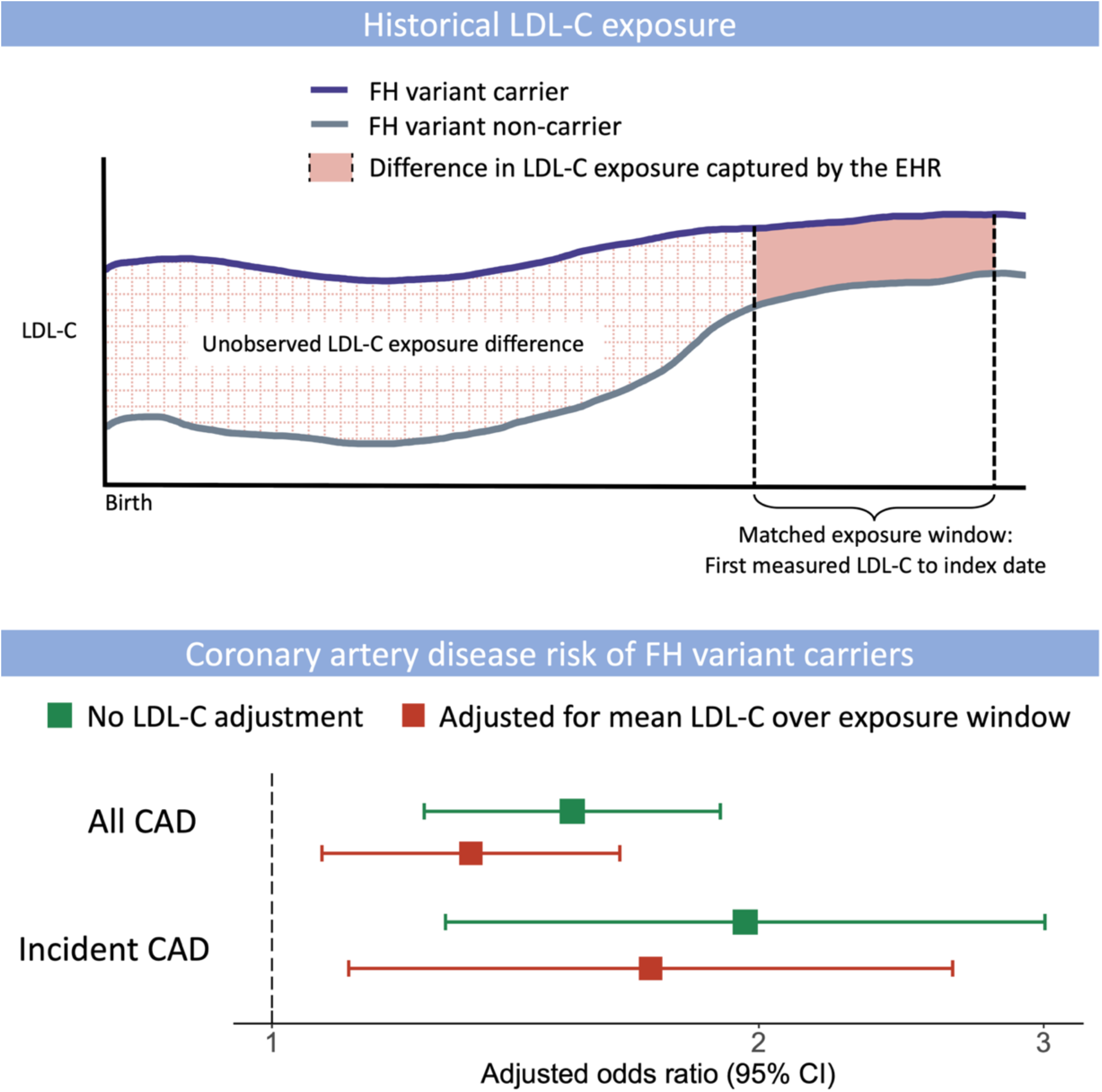

**Take Home Figure. Coronary Artery Disease Risk Associated with Familial Hypercholesterolemia Genetic Variants After Adjustment for Historical LDL-C Cholesterol exposure**. Proposed model of coronary artery disease (CAD) risk associated with familial hypercholesterolemia (FH) genetic variants, independent of measured historical low-density lipoprotein cholesterol (LDL-C) exposure. Longitudinal LDL-C measurements in adults capture some but not all of the increased CAD risk for FH carriers. The residual risk likely reflects differences in lifetime LDL-C exposure beyond what can be measured as part of routine clinical care.

