## Supplementary material for "Coronary Artery Disease Risk of Familial Hypercholesterolemia Genetic Variants Independent of Historical Cholesterol Exposure"

**Title**

**Table S1. Diagnosis and procedure codes used to define cases and controls.**

| Acute MI | | Coronary revascularization | | | Other CAD-related codes | |
|---|---|---|---|---|---|---|
| ICD-9 | ICD-10 | ICD9 | ICD10 | CPT | ICD9 | ICD10 |
| 410 | I21 | 00.66 | 0210 | 92928 | 410 | I20.0 |
| | I22 | 36.0 | 0211 | 92933 | 412 | I21 |
| | | 36.01 | 0212 | 92929 | 411.0 | I22 |
| | | 36.02 | 0213 | 92934 | 411.1 | I23 |
| | | 36.03 | 0270 | 92937 | 411.81 | I24 |
| | | 36.04 | 0271 | 92938 | 411.89 | I24.1 |
| | | 36.05 | 0272 | 92941 | 412 | I25.1 |
| | | 36.06 | 0273 | 92943 | 414.00 | I25.2 |
| | | 36.07 | 02C0 | 92944 | 414.01 | I25.5 |
| | | 36.09 | 02C1 | G0290 | 414.02 | I25.6 |
| | | 36.1 | 02C3 | G0291 | 414.03 | I25.70 |
| | | 36.11 | 02C4 | C9600 | 414.04 | I25.71 |
| | | 36.12 | | C9601 | 414.05 | I25.72 |
| | | 36.13 | | C9602 | 414.2 | I25.73 |
| | | 36.14 | | C9603 | 414.3 | I25.79 |
| | | 36.15 | | C9604 | 414.4 | I25.810 |
| | | 36.16 | | C9605 | 414.8 | I25.82 |
| | | 36.17 | | C9606 | 414.9 | I25.83 |
| | | 36.19 | | C9607 | V45.81 | I25.84 |
| | | 36.2 | | C9608 | V45.82 | I25.89 |
| | | 99.10 | | 92973 | | I25.9 |
| | | | | 92974 | | Z95.1 |
| | | | | 92975 | | Z98.61 |
| | | | | 92977 | | |
| | | | | 92980 | | |
| | | | | 92981 | | |
| | | | | 92982 | | |
| | | | | 92984 | | |
| | | | | 92995 | | |
| | | | | 92996 | | |
| | | | | 33510 | | |
| | | | | 33511 | | |
| | | | | 33512 | | |
| | | | | 33513 | | |
| | | | | 33514 | | |
| | | | | 33516 | | |
| | | | | 33517 | | |
| | | | | 33518 | | |
| | | | | 33519 | | |
| | | | | 33521 | | |
| | | | | 33522 | | |
| | | | | 33523 | | |
| | | | | 33530 | | |
| | | | | 33533 | | |
| | | | | 33534 | | |
| | | | | 33535 | | |
| | | | | 33536 | | |
| | | | | 33572 | | |

**Table S2. ICD-9 codes for risk factors**

| | |
|---|---|
| Diabetes mellitus | 250.00, 250.01, 250.02, 250.03, 250.1, 250.10, 250.11, 250.12, 250.13, 250.2, 250.20, 250.21, 250.22, 250.23, 250.3, 250.30, 250.31, 250.32, 250.33, 250.4, 250.40, 250.41, 250.42, 250.43, 250.5, 250.50, 250.51, 250.52, 250.53, 250.6, 250.60, 250.61, 250.62, 250.63, 250.7, 250.70, 250.71, 250.72, 250.73, 250.1, 250.80, 250.81, 250.82, 250.83, 250.9, 250.90, 250.91, 250.92, 250.93, |
| Hypertension | 401, 401.1, 401.9 |
| Tobacco use | 305.1, 305.10, 305.11, 305.12, 305.13, 649.0, 649.00, 649.01, 649.02, 649.03, 649.04, V15.82 |

### Table S3. Familial hypercholesterolemia genetic variants identified in the MVP population

| Variant (hg19 coordinates) | Locus | FH variant class | | | Number of heterozygotes | | | | | Minor allele frequency | | | | | Assessment |
|---|---|---|---|---|---|---|---|---|---|---|---|---|---|---|---|
| | | CV | PLOF | PPM | AFR | ASN | EUR | HIS | UNK | AFR | ASN | EUR | HIS | UNK | |
| 19:11210962:G:A | LDLR | Y | Y | N | 1 | 1 | 2 | 2 | 0 | 5.7E-06 | 1.1E-04 | 3.1E-06 | 1.5E-05 | 0 | P/LP |
| 19:11210968:G:C | LDLR | Y | N | Y | 0 | 0 | 3 | 0 | 0 | 0 | 0 | 4.7E-06 | 0 | 0 | LP |
| 19:11213381:C:T | LDLR | Y | N | N | 1 | 0 | 29 | 1 | 2 | 5.7E-06 | 0 | 4.6E-05 | 1.5E-05 | 1.3E-04 | LP |
| 19:11213408:T:G | LDLR | Y | N | N | 0 | 0 | 12 | 0 | 1 | 0 | 0 | 2.2E-05 | 0 | 6.5E-05 | P/LP |
| 19:11213441:G:A | LDLR | Y | N | N | 12 | 0 | 12 | 0 | 0 | 6.9E-05 | 0 | 1.9E-05 | 0 | 0 | LP |
| 19:11213445:C:G | LDLR | Y | Y | N | 3 | 0 | 10 | 0 | 0 | 1.7E-05 | 0 | 1.6E-05 | 0 | 0 | P |
| 19:11213450:G:A | LDLR | Y | N | Y | 3 | 0 | 12 | 0 | 0 | 2.3E-05 | 0 | 1.9E-05 | 0 | 0 | P/LP |
| 19:11215908:G:C | LDLR | Y | N | N | 0 | 0 | 11 | 1 | 0 | 0 | 0 | 1.7E-05 | 1.5E-05 | 0 | LP |
| 19:11215944:G:A | LDLR | Y | N | Y | 2 | 0 | 4 | 0 | 0 | 1.1E-05 | 0 | 6.3E-06 | 0 | 0 | LP |
| 19:11216172:G:T | LDLR | Y | N | Y | 1 | 0 | 2 | 2 | 0 | 5.7E-06 | 0 | 3.1E-06 | 4.4E-05 | 0 | LP |
| 19:11216203:C:T | LDLR | Y | N | N | 0 | 1 | 4 | 1 | 0 | 0 | 1.1E-04 | 7.8E-06 | 1.5E-05 | 0 | P/LP |
| 19:11216275:C:A | LDLR | Y | Y | N | 3 | 1 | 10 | 1 | 0 | 1.7E-05 | 1.1E-04 | 1.6E-05 | 1.5E-05 | 0 | P/LP |
| 19:11217328:G:T | LDLR | Y | N | Y | 3 | 0 | 4 | 0 | 0 | 1.7E-05 | 0 | 6.3E-06 | 0 | 0 | LP |
| 19:11217334:A:G | LDLR | N | N | Y | 0 | 0 | 7 | 1 | 0 | 0 | 0 | 1.1E-05 | 1.5E-05 | 0 | LP |
| 19:11217344:T:A | LDLR | N | N | Y | 1 | 0 | 8 | 1 | 0 | 5.7E-06 | 0 | 1.3E-05 | 1.5E-05 | 0 | LP |
| 19:11218070:A:- | LDLR | N | Y | N | 0 | 0 | 4 | 0 | 0 | 0 | 0 | 6.3E-06 | 0 | 0 | P |
| 19:11218112:G:A | LDLR | N | N | Y | 22 | 0 | 6 | 3 | 0 | 1.4E-04 | 0 | 9.4E-06 | 4.4E-05 | 0 | LP |
| 19:11218160:G:A | LDLR | Y | N | N | 39 | 0 | 4 | 0 | 1 | 2.4E-04 | 0 | 6.3E-06 | 0 | 6.5E-05 | P/LP |
| 19:11221354:G:A | LDLR | Y | N | Y | 25 | 0 | 3 | 1 | 2 | 1.4E-04 | 0 | 4.7E-06 | 1.5E-05 | 1.3E-04 | LP |
| 19:11221442:G:A | LDLR | Y | N | Y | 1 | 0 | 6 | 2 | 1 | 5.7E-06 | 0 | 9.4E-06 | 2.9E-05 | 6.5E-05 | P/LP |
| 19:11223984:G:A* | LDLR | N | N | Y | 13 | 0 | 41 | 4 | 1 | 7.5E-05 | 2.2E-04 | 6.4E-05 | 5.9E-05 | 6.5E-05 | LP |
| 19:11223989:G:A | LDLR | N | N | Y | 1 | 0 | 8 | 0 | 1 | 5.7E-06 | 0 | 1.3E-05 | 0 | 6.5E-05 | LP |
| 19:11224005:C:T | LDLR | N | N | Y | 2 | 0 | 20 | 1 | 0 | 1.1E-05 | 0 | 3.3E-05 | 1.5E-05 | 0 | LP |
| 19:11224008:T:G | LDLR | Y | N | N | 1 | 0 | 1 | 0 | 0 | 5.7E-06 | 0 | 1.6E-06 | 0 | 0 | LP |
| 19:11224014:G:A | LDLR | Y | N | N | 1 | 0 | 19 | 0 | 0 | 5.7E-06 | 0 | 3.0E-05 | 0 | 0 | P/LP |
| 19:11224094:T:C | LDLR | N | N | Y | 0 | 0 | 1 | 1 | 0 | 0 | 0 | 1.6E-06 | 1.5E-05 | 0 | LP |
| 19:11224127:T:A* | LDLR | Y | Y | N | 0 | 0 | 5 | 1 | 0 | 0 | 1.1E-04 | 7.8E-06 | 1.5E-05 | 0 | P/LP |
| 19:11224210:G:A | LDLR | Y | Y | N | 2 | 0 | 6 | 0 | 0 | 1.1E-05 | 0 | 9.4E-06 | 0 | 0 | P/LP |
| 19:11224233:G:A | LDLR | Y | N | N | 1 | 0 | 15 | 1 | 0 | 5.7E-06 | 0 | 2.4E-05 | 1.5E-05 | 0 | P/LP |
| 19:11224319:C:G | LDLR | Y | Y | N | 2 | 0 | 5 | 0 | 0 | 1.1E-05 | 0 | 9.4E-06 | 0 | 0 | P/LP |
| 19:11224328:CT:- | LDLR | N | Y | N | 1 | 0 | 4 | 2 | 0 | 5.7E-06 | 0 | 6.3E-06 | 2.9E-05 | 0 | P |
| 19:11224428:C:T | LDLR | N | N | Y | 0 | 0 | 11 | 0 | 0 | 0 | 0 | 1.7E-05 | 1.5E-05 | 0 | LP |
| 19:11227550:G:A | LDLR | N | N | Y | 25 | 1 | 7 | 2 | 0 | 1.4E-04 | 1.1E-04 | 1.3E-05 | 2.9E-05 | 0 | LP |
| 19:11227559:G:A* | LDLR | Y | Y | N | 0 | 0 | 0 | 0 | 0 | 5.7E-06 | 2.2E-04 | 0 | 0 | 0 | P |
| 19:11227562:T:C | LDLR | Y | N | N | 0 | 0 | 1 | 0 | 0 | 0 | 0 | 1.6E-06 | 0 | 0 | P |
| 19:11227576:C:T | LDLR | N | N | Y | 0 | 2 | 3 | 0 | 1 | 0 | 2.2E-04 | 4.7E-06 | 0 | 6.5E-05 | LP |
| 19:11227604:G:A | LDLR | Y | N | Y | 0 | 0 | 32 | 2 | 1 | 5.7E-06 | 0 | 5.0E-05 | 2.9E-05 | 6.5E-05 | P/LP |

| | | | | | | | | | | | | | | | |
|---|---|---|---|---|---|---|---|---|---|---|---|---|---|---|---|
| 19:11227612:C:T | LDLR | N | N | Y | 3 | 0 | 6 | 0 | 0 | 1.7E-05 | 0 | 9.4E-06 | 0 | 0 | LP |
| 19:11227613:G:A | LDLR | N | N | Y | 0 | 0 | 9 | 1 | 1 | 0 | 0 | 1.4E-05 | 1.5E-05 | 6.5E-05 | LP |
| 19:11230832:C:- | LDLR | N | Y | N | 0 | 0 | 4 | 0 | 0 | 0 | 0 | 6.3E-06 | 0 | 0 | P |
| 19:11230900:C:T | LDLR | Y | Y | N | 1 | 0 | 5 | 3 | 0 | 5.7E-06 | 0 | 7.8E-06 | 4.4E-05 | 0 | P |
| 19:11231057:T:C | LDLR | Y | N | Y | 5 | 0 | 0 | 0 | 0 | 3.4E-05 | 0 | 0 | 0 | 0 | LP |
| 19:11231058:G:A | LDLR | Y | N | Y | 1 | 0 | 9 | 0 | 0 | 5.7E-06 | 0 | 1.4E-05 | 0 | 0 | P/LP |
| 19:11231108:G:A | LDLR | Y | N | Y | 1 | 1 | 1 | 0 | 0 | 5.7E-06 | 1.1E-04 | 1.6E-06 | 0 | 0 | LP |
| 19:11231154:C:T | LDLR | N | N | Y | 52 | 0 | 12 | 7 | 4 | 3.2E-04 | 0 | 1.9E-05 | 1.0E-04 | 2.0E-04 | LP |
| 19:11231199:G:A | LDLR | Y | Y | N | 0 | 0 | 12 | 0 | 0 | 0 | 0 | 1.9E-05 | 0 | 0 | P/LP |
| 19:11238694:C:- | LDLR | N | Y | N | 2 | 1 | 9 | 1 | 0 | 1.1E-05 | 1.1E-04 | 1.4E-05 | 1.5E-05 | 0 | P |
| 19:11240186:C:T | LDLR | N | Y | N | 3 | 0 | 20 | 0 | 1 | 1.7E-05 | 0 | 3.3E-05 | 0 | 0 | P |
| 19:11240274:C:G | LDLR | Y | N | Y | 0 | 0 | 1 | 0 | 0 | 0 | 0 | 1.6E-06 | 0 | 0 | P/LP |
| 19:11240278:G:A | LDLR | N | N | Y | 6 | 1 | 282 | 59 | 3 | 4.7E-05 | 1.1E-04 | 5.0E-04 | 9.1E-04 | 1.5E-04 | LP |
| 19:11241961:A:G | LDLR | N | Y | N | 2 | 0 | 34 | 0 | 1 | 1.1E-05 | 0 | 5.6E-05 | 0 | 6.5E-05 | P |
| 1:55509689:T:A | PCSK9 | Y | N | N | 0 | 0 | 9 | 2 | 0 | 0 | 0 | 1.6E-05 | 2.9E-05 | 0 | P/LP |
| 1:55518073:T:C | PCSK9 | Y | N | N | 0 | 0 | 2 | 0 | 0 | 0 | 0 | 3.1E-06 | 0 | 0 | P |
| 2:21229040:G:A | APOB | Y | N | N | 0 | 1 | 56 | 0 | 2 | 0 | 1.1E-04 | 8.9E-05 | 0 | 1.3E-04 | P |
| 2:21229160:C:T | APOB | N | N | Y | 16 | 1 | 293 | 8 | 6 | 9.2E-05 | 1.1E-04 | 4.7E-04 | 1.2E-04 | 3.9E-04 | LP |

CV = ClinVar; PLOF = predicted loss of function; PPM = predicted pathogenic missense; AFR = non-Hispanic African ancestry; ASN = non-Hispanic Asian ancestry; EUR = non-Hispanic European ancestry; HIS = Hispanic ancestry; UNK = Ancestry could not be classified; P = pathogenic; LP = likely pathogenic. Assessment lists the ClinVar pathogenicity interpretation if the variant was defined using ClinVar. Otherwise, it lists the pathogenicity assessment using the American College of Medical Genetics and Genomics guidelines.[1]

* Variant has zero heterozygotes for an ancestry group despite MAF>0 due to no subjects meeting quality criteria.

**Table S4. The association with CAD of two *APOB* missense variants that were previously found to be associated with severe hypercholesterolemia in the MVP population[2] (OR adjusted for sex, year of birth, ancestry, statin, tobacco use, hypertension, and diabetes).**

| Variant (hg19) | ClinVar Classification | ClinVar submissions supporting pathogenic | Clinvar submissions supporting benign | Clinvar submissions supporting uncertain or conflicting | Number of heterozygotes | OR (95% CI) | P |
|---|---|---|---|---|---|---|---|
| 2:21229160:C:T | Conflicting interpretations of pathogenicity | 16 | 0 | 1 | 324 | 3.4 (2.4-4.7) | 1.7E-13 |
| 2:21229068:G:A | Uncertain significance | 0 | 0 | 8 | 493 | 1.0 (0.7-1.4) | 0.9 |

**Table S5. Characteristics of the standard case-control cohort by cases status, based on data from the full span of the EHR.**

| | Cases | Controls | Neither |
|---|---|---|---|
| Demographics | | | |
| n | 34,932 | 291,408 | 109,606 |
| Male | 34,149 (97.8) | 259,423 (89.0) | 105,384 (96.1) |
| Age at enrollment (years) | 67.0 ± 9.0 | 58.7 ± 14.5 | 68.4 ± 10.5 |
| Ancestry | | | |
| African | 4,939 (14.1) | 59,898 (20.6) | 17,487 (16.0) |
| Asian | 213 (0.6) | 3,672 (1.3) | 591 (0.5) |
| European | 27,185 (77.8) | 197,523 (67.8) | 84,328 (76.9) |
| Hispanic | 2,077 (5.9) | 24,708 (8.5) | 5,918 (5.4) |
| Unclassified | 518 (1.5) | 5,607 (1.9) | 1,282 (1.2) |
| FH variant carrier | 172 (0.5) | 890 (0.3) | 435 (0.4) |
| LDL-C (mmol/L) | | | |
| First ever | 3.3 ± 1.1 | 3.1 ± 1.0 | 3.1 ± 1.1 |
| Maximum ever | 4.8 ± 1.4 | 4.0 ± 1.3 | 4.3 ± 1.4 |
| Mean of all measures | 3.0 ± 0.7 | 3.0 ± 0.8 | 2.9 ± 0.8 |
| Medical History | | | |
| Hypertension | 32,039 (91.7) | 154,700 (53.1) | 90,878 (82.9) |
| Diabetes | 19,176 (54.9) | 66,861 (22.9) | 48,383 (44.1) |
| Tobacco | 15,782 (45.2) | 78.275 (26.9) | 34,605 (31.2) |
| Statin use anytime | 33,856 (96.9) | 152,744 (52.4) | 92,796 (84.7) |
| Coronary artery disease | | | |
| Prevalent cases | 29,300 (83.9) | NA | NA |
| Incident cases | 5,632 (16.1) | NA | NA |
| EHR follow up time (years) | | | |
| since first CAD code | 10.0 ± 5.7 | NA | NA |
| from CAD to enrollment | 7.6 ± 4.9 | NA | NA |
| from enrollment to CAD | 2.0 ± 1.5 | NA | NA |

Values are n (%) or mean ± SD.
FH = familial hypercholesterolemia
CAD = coronary artery disease

### Table S6. CAD logistic regression of the nested case-control cohort, comparing adjustments for LDL-C metrics

| | No LDL-C adjustment | | | | Adjusted for first LDL-C | | | | Adjusted for max LDL-C | | | | Adjusted for mean LDL-C | | | |
|---|---|---|---|---|---|---|---|---|---|---|---|---|---|---|---|---|
| | OR | LCI | UCI | P | OR | LCI | UCI | P | OR | LCI | UCI | P | OR | LCI | UCI | P |
| (Intercept) | 0.03 | 0.00 | 0.95 | 4.7E-02 | 0.09 | 0.00 | 2.61 | 1.6E-01 | 2.15 | 0.08 | 60.43 | 6.5E-01 | 0.92 | 0.03 | 25.68 | 9.6E-01 |
| FH variant carrier | 1.53 | 1.24 | 1.89 | 7.0E-05 | 1.41 | 1.14 | 1.74 | 1.5E-03 | 1.34 | 1.09 | 1.66 | 6.3E-03 | 1.33 | 1.07 | 1.64 | 8.8E-03 |
| Tobacco use | 1.56 | 1.51 | 1.60 | 3.2E-188 | 1.56 | 1.52 | 1.61 | 4.3E-191 | 1.55 | 1.51 | 1.60 | 2.1E-184 | 1.57 | 1.52 | 1.62 | 4.1E-193 |
| Hypertension | 1.62 | 1.56 | 1.67 | 1.5E-164 | 1.65 | 1.60 | 1.71 | 1.3E-178 | 1.65 | 1.60 | 1.71 | 2.2E-178 | 1.68 | 1.62 | 1.74 | 4.2E-189 |
| Diabetes | 1.61 | 1.56 | 1.65 | <1E-200 | 1.69 | 1.64 | 1.75 | <1E-200 | 1.72 | 1.67 | 1.77 | <1E-200 | 1.77 | 1.72 | 1.83 | <1E-200 |
| No. of LDL-C measures | 0.99 | 0.99 | 0.99 | 4.3E-19 | 0.99 | 0.99 | 0.99 | 9.7E-18 | 0.98 | 0.98 | 0.99 | 1.3E-49 | 0.99 | 0.99 | 1.00 | 4.6E-10 |
| Statin prescription | 1.52 | 1.48 | 1.57 | 8.4E-150 | 1.36 | 1.32 | 1.41 | 1.3E-73 | 1.23 | 1.19 | 1.28 | 3.2E-30 | 1.25 | 1.21 | 1.30 | 1.6E-36 |
| Year of birth | 1.00 | 1.00 | 1.00 | 8.8E-01 | 1.00 | 1.00 | 1.00 | 4.7E-01 | 1.00 | 1.00 | 1.00 | 9.6E-03 | 1.00 | 1.00 | 1.00 | 2.5E-02 |
| African ancestry | 0.90 | 0.86 | 0.93 | 3.1E-08 | 0.90 | 0.87 | 0.94 | 1.6E-07 | 0.89 | 0.86 | 0.92 | 3.8E-09 | 0.90 | 0.86 | 0.93 | 5.3E-08 |
| Asian ancestry | 0.95 | 0.80 | 1.13 | 5.8E-01 | 0.96 | 0.80 | 1.14 | 6.2E-01 | 0.95 | 0.79 | 1.13 | 5.3E-01 | 0.96 | 0.80 | 1.14 | 6.3E-01 |
| Hispanic ancestry | 0.97 | 0.91 | 1.03 | 2.7E-01 | 0.97 | 0.92 | 1.03 | 3.5E-01 | 0.97 | 0.91 | 1.02 | 2.4E-01 | 0.97 | 0.92 | 1.03 | 3.4E-01 |
| Unclassified ancestry | 0.97 | 0.87 | 1.09 | 6.4E-01 | 0.97 | 0.87 | 1.09 | 6.3E-01 | 0.96 | 0.86 | 1.08 | 4.8E-01 | 0.97 | 0.86 | 1.08 | 5.7E-01 |
| Female | 1.12 | 1.02 | 1.22 | 1.3E-02 | 1.11 | 1.01 | 1.21 | 2.3E-02 | 1.08 | 0.99 | 1.18 | 8.5E-02 | 1.09 | 0.99 | 1.18 | 6.5E-02 |
| First LDL-C: 2003-2007 | 0.98 | 0.95 | 1.02 | 3.1E-01 | 1.01 | 0.98 | 1.04 | 5.9E-01 | 1.01 | 0.98 | 1.05 | 4.2E-01 | 1.03 | 1.00 | 1.06 | 9.2E-02 |
| First LDL-C: 2008-2012 | 1.06 | 1.02 | 1.11 | 2.8E-03 | 1.11 | 1.07 | 1.16 | 3.1E-07 | 1.12 | 1.07 | 1.16 | 6.5E-08 | 1.14 | 1.09 | 1.19 | 4.0E-10 |
| First LDL-C: 2013-2018 | 1.33 | 1.23 | 1.43 | 3.5E-13 | 1.41 | 1.31 | 1.53 | 8.1E-19 | 1.45 | 1.34 | 1.56 | 4.1E-21 | 1.47 | 1.36 | 1.59 | 9.3E-23 |
| LDL-C (per mmol/L) | NA | NA | NA | NA | 1.15 | 1.14 | 1.17 | 1.9E-90 | 1.18 | 1.16 | 1.19 | 3.4E-148 | 1.27 | 1.25 | 1.29 | 1.2E-165 |

**Table S7. MVP cohort characteristics by sex and FH variant carrier status, based on data from the full span of the EHR.**

| | Male | | Female | |
|---|---|---|---|---|
| | **FH variant non-carrier** | **FH variant carrier** | **FH variant non-carrier** | **FH variant carrier** |
| Demographics | | | | |
| n | 397,584 | 1,372 | 36,865 | 125 |
| Age at enrollment (years) | 62.8 ± 13.5 | 64.1 ± 14.0 | 50.5 ± 3.5) | 51.7 ± 14.5 |
| Ancestry | | | | |
| African | 71,249 (17.9) | 225 (16.4) | 10,817 (29.3) | 33 (26.4) |
| Asian | 4,127 (1.0) | 10 (0.7) | 338 (0.9) | 1 (0.8) |
| European | 286,116 (72.0) | 1,018 (74.2) | 21,825 (59.2) | 77 (61.6) |
| Hispanic | 29,675 (7.5) | 98 (7.1) | 2,918 (7.9) | 12 (9.6) |
| Unclassified | 6,417 (1.6) | 21 (1.5) | 967 (2.6) | 2 (1.6) |
| Lipid Data | | | | |
| Age at first LDL-C (years) | 55.9 ± 12.8 | 56.8 ± 13.1 | 44.3 ± 12.3 | 45.2 ± 13.4 |
| LDL-C (mmol/L) | | | | |
| First | 3.1 ± 1.0 | 3.8 ± 1.4 | 3.1 ± 1.0 | 4.1 ± 1.6 |
| Maximum | 4.1 ± 1.3 | 5.2 ± 1.9 | 4.2 ± 1.5 | 5.5 ± 2.0 |
| Mean | 3.0 ± 0.8 | 3.6 ± 1.1 | 3.1 ± 0.8 | 4.0 ± 1.2 |
| Medical History | | | | |
| CAD case | 33,986 (8.5) | 163 (11.9) | 774 (2.1) | 9 (7.2) |
| Hypertension | 261,161 (65.7) | 909 (66.3) | 15,510 (42.1) | 48 (38.4) |
| Diabetes | 127,396 (32.0) | 448 (32.7) | 6,561 (17.8) | 21 (16.8) |
| Tobacco | 118,974 (29.9) | 367 (26.7) | 9,287 (25.2) | 34 (27.2) |
| Statin use | 262,067 (65.9) | 1,043 (76.0) | 16,210 (44.0) | 76 (60.8) |

Values are n (%) or mean ± SD.
FH = familial hypercholesterolemia
CAD = coronary artery disease

**Table S8. MVP cohort characteristics by ancestry and FH variant carrier status, based on data from the full span of the EHR.**

| | African ancestry | | European ancestry | |
|---|---|---|---|---|
| | FH variant non-carrier | FH variant carrier | FH variant non-carrier | FH variant carrier |
| Demographics | | | | |
| n | 82,066 | 258 | 307,941 | 1,095 |
| Age at enrollment (years) | 57.6 ± 12.2 | 57.1 ± 11.8 | 63.9 ± 13.5 | 65.5 ± 13.9 |
| Male | 71,249 (86.9) | 225 (87.2) | 286,116 (92.9) | 1,018 (93.0) |
| Lipid Data | | | | |
| Age at first LDL-C (years) | 50.4 ± 11.4 | 49.8 ± 11.1 | 63.9 ± 13.5 | 65.5 ± 13.9 |
| LDL-C (mmol/L) | | | | |
| First | 3.1 ± 1.0 | 4.1 ± 1.4 | 3.1 ± 1.0 | 3.7 ± 1.5 |
| Maximum | 4.2 ± 1.5 | 5.6 ± 2.0 | 4.1 ± 1.3 | 5.1 ± 1.9 |
| Mean | 3.0 ± 0.8 | 3.9 ± 1.1 | 3.0 ± 0.8 | 3.6 ± 1.1 |
| Medical History | | | | |
| CAD case | 4,908 (6.0) | 31 (12.0) | 27,059 (8.8) | 126 (11.5) |
| Hypertension | 57,465 (70.0) | 181 (70.2) | 195,965 (63.6) | 706 (64.5) |
| Diabetes | 29,453 (35.9) | 98 (38.0) | 90,419 (29.4) | 232 (21.2) |
| Tobacco | 29,215 (35.6) | 98 (38.0) | 88,805 (28.8) | 278 (25.4) |
| Statin use | 51,755 (63.1) | 200 (77.5) | 201,458 (65.4) | 827 (75.5) |

Values are n (%) or mean ± SD.
FH = familial hypercholesterolemia
CAD = coronary artery disease

**Table S9. CAD logistic regression of the nested case-control cohort restricted to subjects with a minimum of 5 LDL-C measures spanning at least 5 years prior to the index date**

| | No LDL-C adjustment | | | | Adjusted for first LDL-C | | | | Adjusted for max LDL-C | | | | Adjusted for mean LDL-C | | | |
|---|---|---|---|---|---|---|---|---|---|---|---|---|---|---|---|---|
| | **OR** | **LCI** | **UCI** | **P** | **OR** | **LCI** | **UCI** | **P** | **OR** | **LCI** | **UCI** | **P** | **OR** | **LCI** | **UCI** | **P** |
| (Intercept) | 0.07 | 0.00 | 10.94 | 3.0E-01 | 0.11 | 0.00 | 16.25 | 3.8E-01 | 10.89 | 0.07 | 1727.57 | 3.6E-01 | 6.96 | 0.04 | 1097.38 | 4.5E-01 |
| FH variant carrier | 1.83 | 1.35 | 2.48 | 1.0E-04 | 1.72 | 1.27 | 2.33 | 4.9E-04 | 1.58 | 1.16 | 2.15 | 3.4E-03 | 1.56 | 1.15 | 2.12 | 4.4E-03 |
| Tobacco use | 1.57 | 1.50 | 1.64 | 3.4E-90 | 1.58 | 1.51 | 1.65 | 2.8E-91 | 1.57 | 1.50 | 1.64 | 1.9E-89 | 1.58 | 1.51 | 1.65 | 5.7E-93 |
| Hypertension | 1.81 | 1.70 | 1.93 | 3.2E-75 | 1.83 | 1.72 | 1.95 | 3.3E-78 | 1.85 | 1.74 | 1.97 | 4.9E-81 | 1.88 | 1.77 | 2.01 | 3.3E-85 |
| Diabetes | 1.52 | 1.45 | 1.59 | 3.8E-73 | 1.57 | 1.50 | 1.65 | 4.3E-83 | 1.63 | 1.56 | 1.71 | 1.2E-95 | 1.70 | 1.63 | 1.79 | 1.3E-108 |
| No. of LDL-C measures | 1.00 | 1.00 | 1.01 | 1.0E-02 | 1.00 | 1.00 | 1.01 | 3.5E-02 | 1.00 | 0.99 | 1.00 | 1.3E-01 | 1.00 | 1.00 | 1.01 | 2.9E-02 |
| Statin prescription | 1.55 | 1.46 | 1.63 | 2.3E-54 | 1.42 | 1.34 | 1.50 | 3.1E-32 | 1.21 | 1.14 | 1.29 | 8.2E-10 | 1.22 | 1.14 | 1.29 | 4.6E-10 |
| Year of birth | 1.00 | 1.00 | 1.00 | 7.3E-01 | 1.00 | 1.00 | 1.00 | 5.4E-01 | 1.00 | 0.99 | 1.00 | 1.3E-02 | 1.00 | 0.99 | 1.00 | 1.5E-02 |
| African ancestry | 0.89 | 0.84 | 0.94 | 6.8E-05 | 0.89 | 0.84 | 0.95 | 1.0E-04 | 0.88 | 0.83 | 0.93 | 8.3E-06 | 0.88 | 0.84 | 0.94 | 2.5E-05 |
| Asian ancestry | 0.97 | 0.76 | 1.24 | 8.0E-01 | 0.97 | 0.76 | 1.25 | 8.3E-01 | 0.97 | 0.76 | 1.24 | 7.9E-01 | 0.99 | 0.77 | 1.27 | 9.4E-01 |
| Hispanic ancestry | 0.94 | 0.86 | 1.03 | 2.0E-01 | 0.95 | 0.87 | 1.04 | 2.6E-01 | 0.94 | 0.86 | 1.03 | 1.6E-01 | 0.95 | 0.87 | 1.04 | 2.4E-01 |
| Unclassified ancestry | 0.99 | 0.84 | 1.18 | 9.3E-01 | 0.99 | 0.84 | 1.18 | 9.5E-01 | 0.98 | 0.82 | 1.16 | 8.0E-01 | 0.98 | 0.83 | 1.17 | 8.6E-01 |
| Female | 1.15 | 1.02 | 1.30 | 2.2E-02 | 1.15 | 1.02 | 1.30 | 2.7E-02 | 1.10 | 0.97 | 1.24 | 1.4E-01 | 1.10 | 0.97 | 1.24 | 1.3E-01 |
| First LDL-C: 2003-2007 | 1.06 | 1.01 | 1.11 | 1.9E-02 | 1.07 | 1.02 | 1.12 | 4.8E-03 | 1.08 | 1.03 | 1.13 | 9.2E-04 | 1.09 | 1.04 | 1.15 | 1.5E-04 |
| First LDL-C: 2008-2012 | 1.18 | 1.09 | 1.28 | 3.9E-05 | 1.21 | 1.12 | 1.31 | 3.2E-06 | 1.22 | 1.13 | 1.33 | 7.0E-07 | 1.24 | 1.15 | 1.34 | 1.3E-07 |
| First LDL-C: 2013-2018 | 4.43 | 0.88 | 22.29 | 7.1E-02 | 4.27 | 0.85 | 21.36 | 7.7E-02 | 4.05 | 0.80 | 20.48 | 9.0E-02 | 4.30 | 0.86 | 21.61 | 7.6E-02 |
| LDL-C (per mmol/L) | NA | NA | NA | NA | 1.10 | 1.08 | 1.13 | 2.3E-19 | 1.18 | 1.15 | 1.20 | 8.0E-68 | 1.31 | 1.27 | 1.35 | 1.6E-68 |

**Table S10. CAD logistic regression of the nested case-control cohort restricted to subjects with a minimum of 10 LDL-C measures spanning at least 10 years prior to the index date**

| | No LDL-C adjustment | | | | Adjusted for first LDL-C | | | | Adjusted for max LDL-C | | | | Adjusted for mean LDL-C | | | |
|---|---|---|---|---|---|---|---|---|---|---|---|---|---|---|---|---|
| | OR | LCI | UCI | P | OR | LCI | UCI | P | OR | LCI | UCI | P | OR | LCI | UCI | P |
| (Intercept) | 3.82 | 0.00 | 1.4E+04 | 7.5E-01 | 3.95 | 0.00 | 1.4E+04 | 7.4E-01 | 176.62 | 0.05 | 6.7E+05 | 2.2E-01 | 112.09 | 0.03 | 4.3E+05 | 2.6E-01 |
| FH variant carrier | 2.16 | 1.39 | 3.37 | 6.3E-04 | 2.12 | 1.36 | 3.29 | 9.1E-04 | 1.99 | 1.28 | 3.11 | 2.3E-03 | 2.01 | 1.29 | 3.13 | 2.1E-03 |
| Tobacco use | 1.53 | 1.43 | 1.65 | 3.3E-31 | 1.54 | 1.43 | 1.65 | 2.8E-31 | 1.53 | 1.42 | 1.65 | 7.7E-31 | 1.54 | 1.43 | 1.66 | 1.3E-31 |
| Hypertension | 1.93 | 1.71 | 2.16 | 5.5E-28 | 1.93 | 1.72 | 2.18 | 2.3E-28 | 1.96 | 1.74 | 2.20 | 2.7E-29 | 1.98 | 1.76 | 2.23 | 2.4E-30 |
| Diabetes | 1.55 | 1.44 | 1.67 | 6.7E-31 | 1.57 | 1.46 | 1.69 | 3.4E-32 | 1.62 | 1.51 | 1.75 | 7.8E-37 | 1.68 | 1.55 | 1.81 | 1.2E-39 |
| No. of LDL-C measures | 1.01 | 1.00 | 1.01 | 3.4E-03 | 1.01 | 1.00 | 1.01 | 6.0E-03 | 1.00 | 1.00 | 1.01 | 3.6E-01 | 1.01 | 1.00 | 1.01 | 1.8E-02 |
| Statin prescription | 1.54 | 1.39 | 1.71 | 6.4E-17 | 1.48 | 1.33 | 1.64 | 7.1E-13 | 1.28 | 1.14 | 1.43 | 1.8E-05 | 1.29 | 1.16 | 1.45 | 6.4E-06 |
| Year of birth | 1.00 | 0.99 | 1.00 | 2.2E-01 | 1.00 | 0.99 | 1.00 | 2.1E-01 | 1.00 | 0.99 | 1.00 | 2.8E-02 | 1.00 | 0.99 | 1.00 | 3.2E-02 |
| African ancestry | 0.89 | 0.81 | 0.98 | 1.6E-02 | 0.90 | 0.82 | 0.98 | 1.8E-02 | 0.88 | 0.81 | 0.97 | 8.1E-03 | 0.89 | 0.81 | 0.97 | 1.1E-02 |
| Asian ancestry | 0.98 | 0.69 | 1.40 | 9.2E-01 | 0.98 | 0.69 | 1.41 | 9.2E-01 | 0.98 | 0.69 | 1.41 | 9.3E-01 | 1.00 | 0.70 | 1.43 | 1.0E+00 |
| Hispanic ancestry | 0.93 | 0.81 | 1.08 | 3.4E-01 | 0.94 | 0.81 | 1.08 | 3.8E-01 | 0.93 | 0.81 | 1.08 | 3.6E-01 | 0.94 | 0.82 | 1.09 | 4.4E-01 |
| Unclassified ancestry | 0.99 | 0.75 | 1.32 | 9.5E-01 | 0.99 | 0.74 | 1.31 | 9.3E-01 | 0.98 | 0.74 | 1.30 | 8.8E-01 | 0.99 | 0.74 | 1.31 | 9.2E-01 |
| Female | 1.17 | 0.97 | 1.42 | 1.1E-01 | 1.17 | 0.96 | 1.41 | 1.1E-01 | 1.12 | 0.93 | 1.36 | 2.4E-01 | 1.13 | 0.93 | 1.36 | 2.3E-01 |
| First LDL-C: 2003-2007 | 1.10 | 1.02 | 1.19 | 1.8E-02 | 1.11 | 1.02 | 1.20 | 1.3E-02 | 1.12 | 1.03 | 1.21 | 5.9E-03 | 1.13 | 1.04 | 1.22 | 3.8E-03 |
| First LDL-C: 2008-2012 | 3.40 | 0.70 | 16.52 | 1.3E-01 | 3.52 | 0.72 | 17.08 | 1.2E-01 | 3.62 | 0.74 | 17.61 | 1.1E-01 | 3.83 | 0.79 | 18.62 | 9.6E-02 |
| LDL-C (per mmol/L) | NA | NA | NA | NA | 1.05 | 1.01 | 1.08 | 9.6E-03 | 1.13 | 1.09 | 1.16 | 8.0E-16 | 1.22 | 1.16 | 1.29 | 3.0E-14 |

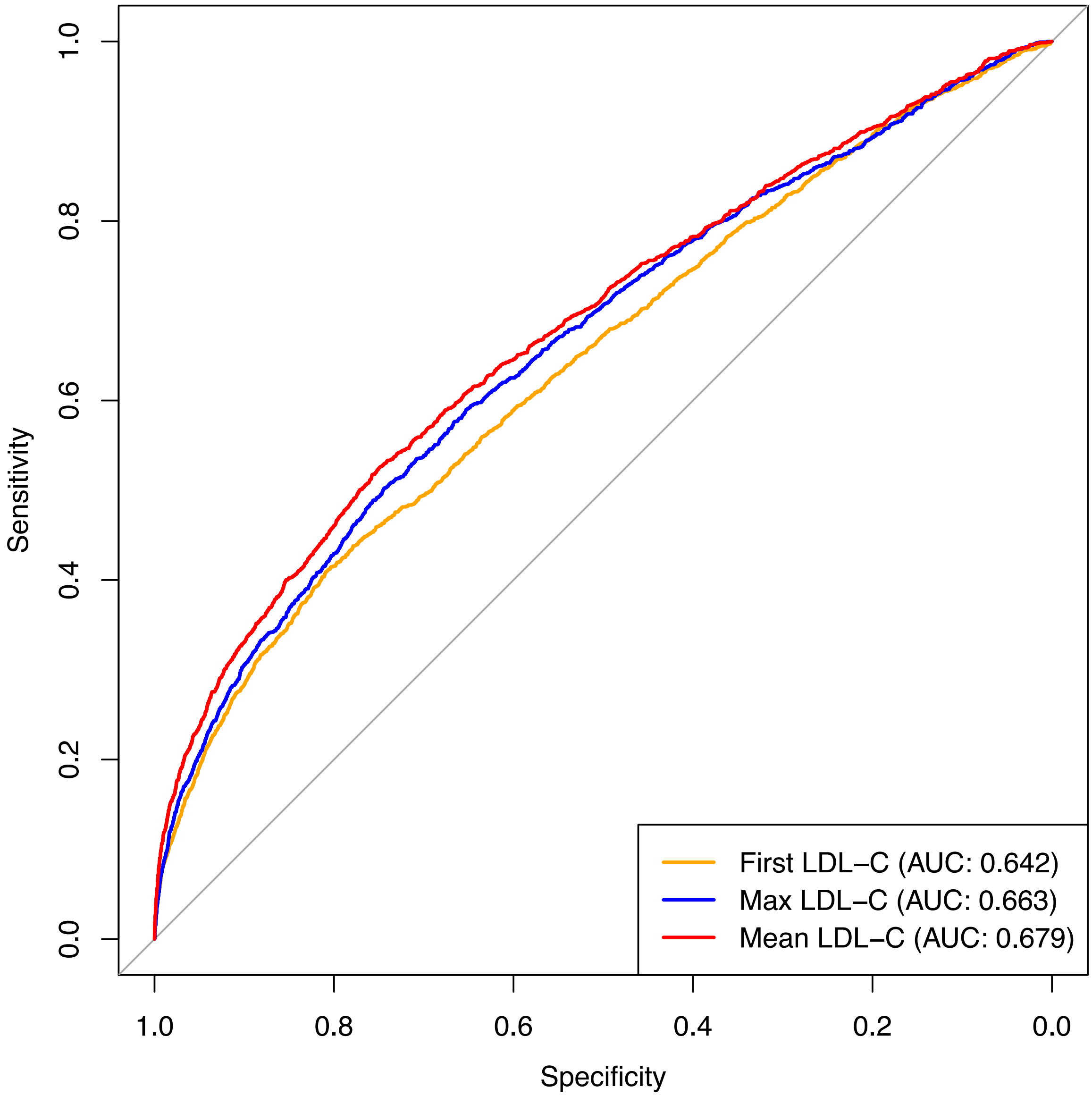


**Figure S1. Receiver operating characteristic curve for predicting the presence of an FH variant using LDL-C and age of measurement.** For Mean LDL-C, the age of each measurement was used to calculate a mean age.

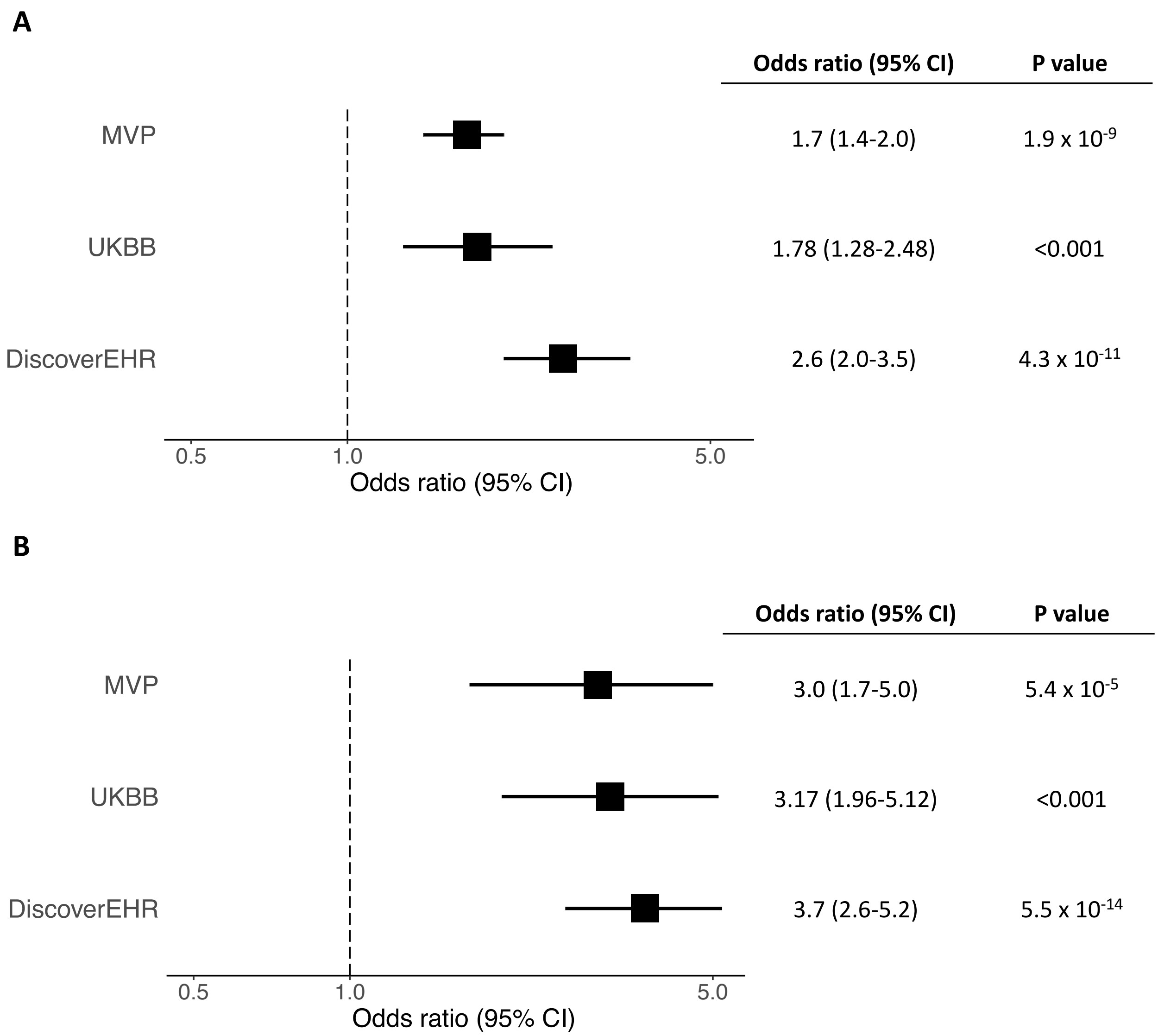


**Figure S2. Risk of all CAD (A) and premature CAD (B) associated with FH variants in MVP compared to the UK Biobank (UKBB) and the MyCode Community Health Initiative of the Geisinger Health System (DiscoverEHR).** The odds ratios in MVP were calculated using the standard case-control analysis. The odds ratios and p values for UKBB and DiscoverEHR are as reported in those respective publications.[3,4] For MVP and DiscoverEHR, the outcome is CAD. For UKBB, the outcome is CAD and stroke. For MVP and DiscoverEHR, premature CAD was defined as males ≤ 55 and females ≤ 65. For the UKBB study, premature CAD/stroke was defined as males and females ≤ 55.

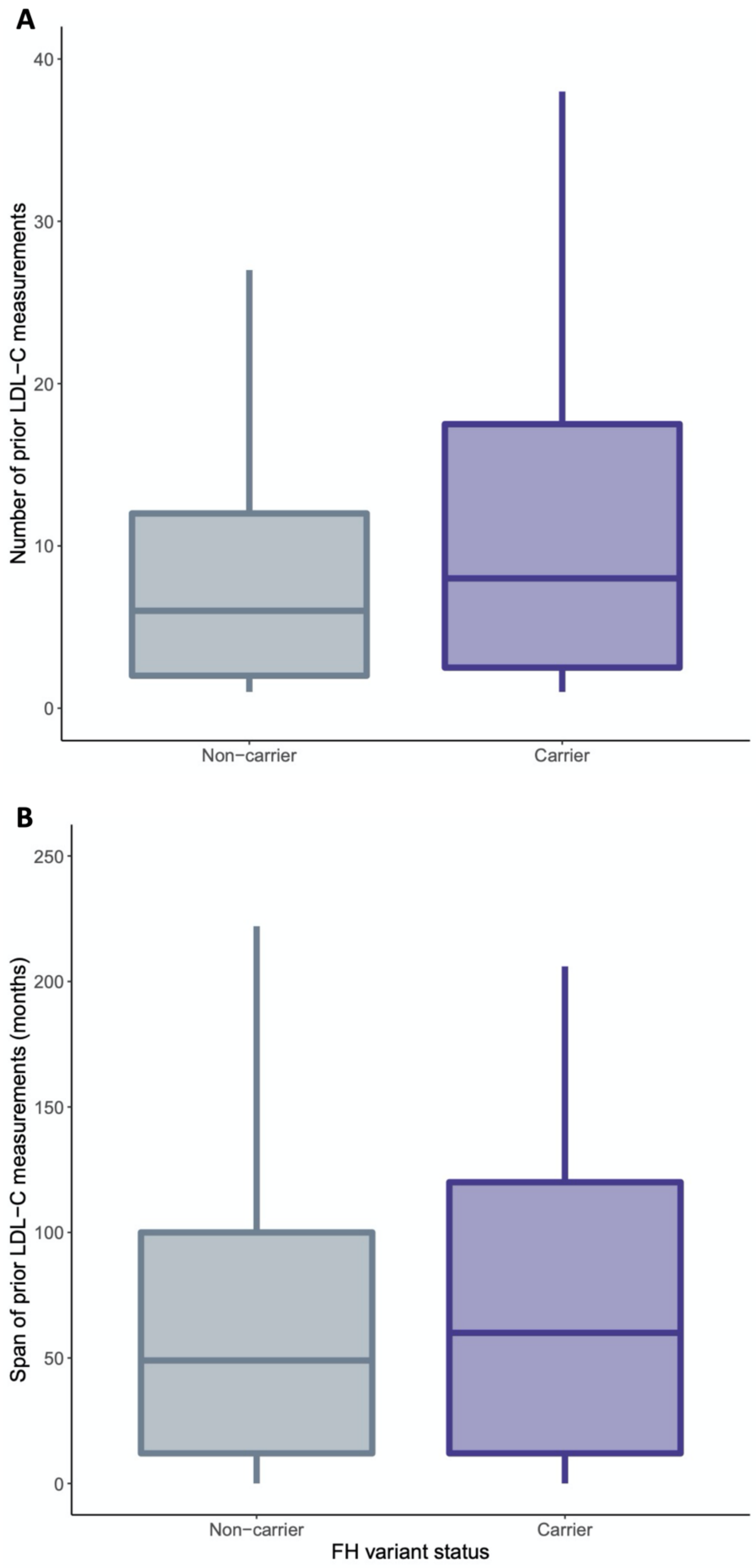


**Figure S3. Extent of LDL-C data among CAD cases prior to the diagnosis of CAD.** Boxplots showing the number of low-density lipoprotein (LDL-C) measurements and the span of those measurements (months) among coronary artery disease (CAD) cases prior to diagnosis, stratified by familial hypercholesterolemia (FH) carrier status.

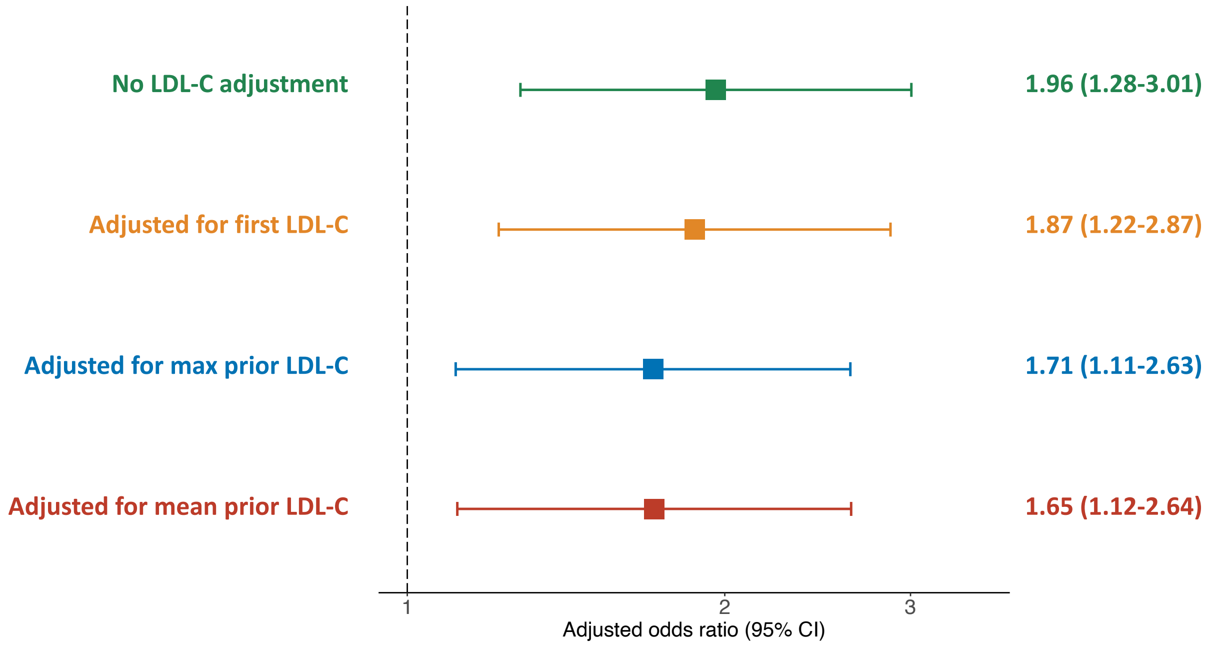


**Figure S4. Association between FH variants and incident CAD with adjustments for historical LDL-C exposure.** Risk of incident coronary artery disease (CAD) associated with familial hypercholesterolemia (FH) genetic variants (5,449 cases and 54,490 matched controls). Adjustment for the first, maximum observed, or mean low-density lipoprotein cholesterol (LDL-C) prior to the index date does not fully attenuate the risk associated with FH. Odds ratios were estimated using logistic regression, adjusting for the nested case-control matching factors, tobacco use, hypertension, diabetes, statin prescription, and number of LDL-C measurements.

**Supplementary material references**


1. Richards S, Aziz N, Bale S, Bick D, Das S, Gastier-Foster J, Grody WW, Hegde M, Lyon E, Spector E, Voelkerding K, Rehm HL, ACMG Laboratory Quality Assurance Committee. Standards and guidelines for the interpretation of sequence variants: a joint consensus recommendation of the American College of Medical Genetics and Genomics and the Association for Molecular Pathology. *Genet Med Off J Am Coll Med Genet* 2015;**17**:405–424.

2. Sun YV, Damrauer SM, Hui Q, Assimes TL, Ho Y-L, Natarajan P, Klarin D, Huang J, Lynch J, DuVall SL, Pyarajan S, Honerlaw JP, Gaziano JM, Cho K, Rader DJ, O'Donnell CJ, Tsao PS, Wilson PWF. Effects of Genetic Variants Associated with Familial Hypercholesterolemia on Low-Density Lipoprotein-Cholesterol Levels and Cardiovascular Outcomes in the Million Veteran Program. *Circ Genomic Precis Med* 2018;**11**.

3. Trinder M, Francis GA, Brunham LR. Association of Monogenic vs Polygenic Hypercholesterolemia With Risk of Atherosclerotic Cardiovascular Disease. *JAMA Cardiol* 2020;

4. Abul-Husn NS, Manickam K, Jones LK, Wright EA, Hartzel DN, Gonzaga-Jauregui C, O'Dushlaine C, Leader JB, Lester Kirchner H, Lindbuchler DM, Barr ML, Giovanni MA, Ritchie MD, Overton JD, Reid JG, Metpally RPR, Wardeh AH, Borecki IB, Yancopoulos GD, Baras A, Shuldiner AR, Gottesman O, Ledbetter DH, Carey DJ, Dewey FE, Murray MF. Genetic identification of familial hypercholesterolemia within a single U.S. health care system. *Science* 2016;**354**.
